# Integrative multi-omics analysis of genomic, epigenomic, and metabolomics data leads to new insights for Attention-Deficit/Hyperactivity Disorder

**DOI:** 10.1101/2022.07.21.22277887

**Authors:** Nikki Hubers, Fiona A. Hagenbeek, René Pool, Sébastien Déjean, Amy C. Harms, Peter J. Roetman, Catharina E. M. van Beijsterveldt, Vassilios Fanos, Erik A. Ehli, Robert R. J. M. Vermeiren, Meike Bartels, Jouke Jan Hottenga, Thomas Hankemeier, Jenny van Dongen, Dorret I. Boomsma

**Affiliations:** Department of Biological Psychology, Vrije Universiteit Amsterdam, Amsterdam, the Netherlands; Amsterdam Reproduction & Development (AR&D) research institute, Amsterdam, the Netherlands; Amsterdam Public Health research institute, Amsterdam, the Netherlands; Toulouse Mathematics Institute, UMR 5219, University of Toulouse, CNRS, Toulouse, France; Division of Analytical Biosciences, Leiden Academic Center for Drug Research, Leiden University, Leiden, the Netherlands; The Netherlands Metabolomics Centre, Leiden, The Netherlands; LUMC-Curium, Department of Child and Adolescent Psychiatry, Leiden University Medical Center, Leiden, the Netherlands; Department of Surgical Sciences, University of Cagliari and Neonatal Intensive Care Unit, Cagliari, Italy; Avera Institute for Human Genetics, Sioux Falls, South Dakota, USA; Youz, Parnassia Group, the Hague, the Netherlands

**Keywords:** ADHD, multi-omics, polygenic scores, genetic nurture, DNA methylation, metabolites.

## Abstract

The evolving field of multi-omics combines data and provides methods for simultaneous analysis across several omics levels. Here, we integrated genomics (transmitted and non-transmitted polygenic scores (PGS)), epigenomics and metabolomics data in a multi-omics framework to identify biomarkers for ADHD and investigated the connections among the three omics levels. We first trained single- and next multi-omics models to differentiate between cases and controls in 596 twins (cases=14.8%) from the Netherlands Twin Register (NTR) demonstrating reasonable in-sample prediction through cross-validation. The multi-omics model selected 30 PGSs, 143 CpGs, and 90 metabolites. We confirmed previous associations of ADHD with glucocorticoid exposure and the transmembrane protein family *TMEM*, show that the DNA methylation of the *MAD1L1* gene associated with ADHD has a relation with parental smoking behavior, and present novel findings including associations between indirect genetic effects and CpGs of the *STAP2* gene. Out-of-sample prediction in NTR participants (N=258, cases=14.3%) and in a clinical sample (N=145, cases=51%) did not perform well (range misclassification was [0.40, 0.57]). The results highlighted connections between omics levels, with the strongest connections between non-transmitted PGS, CpGs, and amino acid levels and show that multi-omics designs considering interrelated omics levels can help unravel the complex biology underlying ADHD.

## Introduction

Attention-Deficit/Hyperactivity Disorder (ADHD) in children is a highly heritable (75 to 91%) complex neurodevelopmental disorder that is characterized by high levels of hyperactivity, inattention, and impulsiveness (Roehr, 2013). The prevalence of ADHD in children is 5-8%, and a majority of these children continue to display symptoms in adulthood (Derks et al., 2008; Faraone et al., 2018; Levy et al., 1997; Luo et al., 2019). Persons with ADHD are more likely to be extroverted and entrepreneurs but also have an increased lifetime risk of harmful outcomes (Faraone et al., 2021; Martel et al., 2010). Multiple psychiatric conditions are comorbid with ADHD, including borderline personality disorder, anxiety, and conduct/oppositional defiant disorder, resulting in challenges with the diagnosis, referral, and treatment of ADHD (Caye et al., 2019; Demontis et al., 2019; Distel et al., 2011; Joseph et al., 2015; Reale et al., 2017).

Several single-omics studies involving genomics, epigenomics, and metabolomics have been performed to unravel the etiology of ADHD. Large (*N* = 55,374) Genome-Wide Association (GWA) studies initially identified 12 (Demontis et al., 2019) and recently 27 genome-wide significantly associated loci for ADHD (Demontis et al., 2023). Demontis et al. (2019) compared these results with those for continuous measures of ADHD symptoms in the general population (Middeldorp et al., 2016), concluding that the same genetic variants that give rise to an ADHD diagnosis also affect inattention and impulsivity in the general population.

The biological basis of ADHD has also been explored in Epigenome-Wide Association (EWA) and metabolomics studies. Epigenomics constitutes a link between genetic and environmental exposures through changes in gene expression with processes such as DNA methylation (Barros & Offenbacher, 2009). Methylation level variation is associated with exposures such as stressful life events, nutritional factors, and toxins, and is partly due to genetic factors (Moore et al., 2012). Interestingly, it has been hypothesized that the decrease in hyperactivity with age might be due to epigenetic factors regulating the expression of genes involved in hyperactivity (Elia et al., 2012). EWA studies that measured DNA methylation in cord blood at birth, in saliva from older children, or in blood from adults start to provide evidence for DNA methylation sites associated with ADHD symptom trajectories and ADHD clinical diagnosis in children and adults (Goodman et al., 2020; Mooney et al., 2020; Neumann et al., 2020; Rovira, Sanchez-Mora, et al., 2020; Walton, 2019). EWA studies based on blood samples from adolescents and adults identified multiple new candidate loci involved with immune and neuronal functioning associated with adult ADHD symptoms in population-based samples (van Dongen et al., 2019) and loci involved in cholesterol signaling in a comparison of participants with persistent ADHD and controls (Meijer et al., 2020).

Metabolomics studies utilize a profile of small molecules derived from cellular metabolism (X. Liu & Locasale, 2017). The first studies of metabolomics and ADHD identified multiple metabolites associated with ADHD in plasma (Wang et al., 2020). For instance, metabolites involved in oxidative stress pathway(s) were positively associated with ADHD in children. Meta-analyses showed that across all ages, proteins involved in dopamine metabolism and the fatty acid docosahexaenoic acid (DHA) in different tissues were negatively associated with ADHD (Bonvicini et al., 2016, 2018). However, sample sizes of the metabolomics studies concerning ADHD are relatively small and it is likely that other metabolites and biological pathways influencing ADHD are still to be identified (Bonvicini et al., 2018).

The growing numbers of single-omics studies for ADHD indicate that large cohorts increasingly collect information across omics levels, including genomics, epigenomics, and metabolomics in relation to health and behavioral outcomes. Single-omics analyses contribute to an understanding of the etiologies for the complex outcomes but fail to include the interactions between the different omics levels. Associations and correlations across omics levels have been reported between the genome and epigenome (Min et al., 2021; van Dongen et al., 2016), the genome and the metabolome (Hagenbeek, Pool, et al., 2020; Hagenbeek, van Dongen, Pool, Harms, et al., 2022)and the epigenome and the metabolome (Gomez-Alonso et al., 2021). Thus, a next step is to combine multiple omics levels into multi-omics approaches to provide a basic understanding of how different omics levels are associated with specific phenotypes such as ADHD (Hasin et al., 2017), but also how they are interconnected. To optimally analyze multi-omics data in association and etiological studies, dedicated statistical treatment of simultaneous omics influences is required (Durufle et al., 2020), where simultaneous modeling complements single omics approaches. Such innovative multi-omics analyses may result in novel insights and uncover new biological pathways underlying traits and diseases (Rajasundaram & Selbig, 2016). For example, in cancer research, multi-omics findings hold promise to be one of the keys leading to improved personalized medicine by identifying disease markers (Chakraborty et al., 2018).

We applied an integrative multi-omics approach to elucidate biological mechanisms underlying ADHD and identify potential biomarkers. We integrated genetic data comprising genome-wide methylation data, and urinary metabolomic data with transmitted and non-transmitted polygenic scores (PGSs) for 15 traits, which are genetically correlated with ADHD. Here the PGS based on alleles transmitted from parents to offspring represent the offspring’s own genetic liability, whereas the non-transmitted PGS represent the environmental influences that are associated with the parental with alleles that were not transmitted (e.g., genetic nurture; Kong et al., 2018). The study included 854 twins (N cases = 125, N controls = 729, mean age = 9.5 years (SD = 1.9), % female = 50.7, 359 complete twin pairs) originating from the Netherlands Twin Register (NTR; Ligthart et al., 2019) and 145 non-twin children from a youth psychiatry clinic. We included non-transmitted PGSs to explore the environmental influences that children are exposed to that are created by the parents’ genetics. To our knowledge, this is one of the first studies to include non-transmitted PGSs in a multi-omics design.

## Materials and Methods

### Study population and procedures

Participants were selected from the Young Netherlands Twin Register (YNTR; van Beijsterveldt et al., 2013) and from a youth psychiatry clinic (LUMC-Curium, the Netherlands), as part of the ACTION (Aggression in Children: Unraveling gene-environment interplay to inform Treatment and InterventiON strategies) Biomarker Study (Bartels et al., 2018; Hagenbeek, et al., 2020). The ACTION project collected first-morning urine samples and buccal-cell swabs from 1494 twins (747 complete pairs) and 189 children from a clinical cohort with standardized protocols (see http://www.action-euproject.eu/content/data-protocols). The first-morning urine samples were stored in a freezer at the child’s home (*T* ≈ −18^◦^C) and were transported to the lab in a mobile freezer unit at *T* = −18^◦^C. In the lab, urine samples were stored at *T* = −80^◦^C until further processing. The buccal swabs were collected on two consecutive days: twice in the morning (before breakfast) and twice in the evening (before dinner). In the twin cohort, buccal-cell swabs were also collected from the parents and siblings of the twins.

ADHD status in the twin cohort was based on sex- and age-specific T-scores from the mother-rated ADHD DSM-oriented scale of the ASEBA-CBCL as completed around the time of biological sample collection. T-scores were calculated in the entire NTR for children with CBCL data available at 9 to 10 years of age (N = 23,858). Twins with ADHD CBCL T-scores of 65 or higher were classified as cases (N = 125) and twins with T-scores below 65 as controls (N = 729). In the LUMC-Curium cohort, parents (90% mothers, 10% fathers) completed a CBCL as part of a standardized clinical assessment a maximum of 6 months before or after biological sample collection. T-scores were calculated via CBCL software. Children with T-scores of 70 or higher were classified as cases (N = 74), children with T-scores below 65 as controls (N = 71), children with T-scores between 65 and 69 were excluded (N = 40).

NTR and LUMC-Curium participants were excluded if no ADHD status at the time of biological sample collection was available (N = 209), if they were assigned a control status while their co-twin was assigned a case status for ADHD (*N* = 51), if the collected urine sample was not the first-morning urine (e.g., parent-reported time of urine collection was after 12:00 in the afternoon, *N* = 13), or if any of the omics data were not available because of poor quality (N = 367). From the twin cohort, 854 out of 1494 participants, and from the clinical cohort, 185 participants out of 189 had complete phenotype and omics data across all omics levels (genomics, epigenomics, and metabolomics).

## Genotyping and polygenic scores

### Genotyping and imputation

Genotyping in the NTR and LUMC-Curium cohorts was performed simultaneously on Affymetrix AXIOM or Illumina GSA arrays (Beck et al., 2019; Ehli et al., 2017), and genome-wide SNP data were available for 3334 participants, including 1702 parents and siblings of twins (AXIOM = 909, GSA = 2425). For each genotyping platform, samples were removed based on the following criteria: 1) DNA sex did not match self-reported sex; 2) Plink heterozygosity F statistic (< -0.10 | > 0.10); and 3) genotyping call rate (< 0.90). Criteria for excluding Single Nucleotide Polymorphisms (SNP) included: 1) minor allele frequency (MAF; <0.005); 2) Hardy-Weinberg Equilibrium (HWE) p-value (< 1×10^-5^), 3) call rate (< 0.95), 4) number of Mendelian errors (> 2); and 5) palindromic AT/GC SNPs (0.4 ≤ MAF ≤ 0.5; Purcell et al., 2007). For each platform, data were aligned with the GoNL reference set (V4; Boomsma et al., 2014). SNPs with an allele frequency difference of larger than 0.10, or mismatching alleles with the reference panel were removed and samples were excluded if DNA Identity by Descent (IBD) state did not match the expected familial relations (PLINKv1.9). Genotypes were then re-aligned, for each platform separately, to the 1000G Phase 3 version 5 reference panel by the PERL based “1000G Imputation preparation and checking” tool v4.3 (https://www.well.ox.ac.uk/~wrayner/tools/). The two platforms were subsequently phased (EAGLE v2.4.1) and imputed (MINIMAC3 v2.0.1) following the Michigan Imputation server protocols (Das et al., 2016; Delaneau & Zagury, 2012; Loh et al., 2016). After imputation, the resulting per-platform chromosomal files were merged into a single best-guess Plink file (PLINKv1.9). Data were available for 3,334 participants: 3,149 NTR participants (1,447 twins, 1,702 parents/siblings of twins), and 185 LUMC-Curium participants.

### Computation of principal components

From the best-guess 1000-genomes imputed data, a complete list of all platform-genotyped SNPs on both AXIOM and Illumina GSA were extracted from the NTR and LUMC-Curium cohorts. This creates a single dataset without large SNP missingness where most genotypic information from both platforms is included. The same SNPs were extracted from the 1000G reference panel genotype data with additional filters for MAF (>0.05), and call rate (>0.98). In the 1000G population alone, the SNPs were then LD pruned (PLINK1.9; --indep 50 5 2) and long-range LD regions were removed (Abdellaoui et al., 2013). Next, the NTR, LUMC-Curium, and 1000 genomes sets were merged for all SNPs that passed the above QC criteria and were present in all sets. Subsequently, twenty principal components were calculated for the 1000 genomes PCs together with our sample to control the polygenic scores for stratification due to ancestry (SMARTPCAv7; Price et al., 2006).

### Transmitted and non-transmitted alleles and calculation of polygenic scores

The NTR cohort included 1,447 twins with genotyped parents, for whom allele transmission could be established based on the imputed best guess data. Allele transmission could not be established in the LUMC-Curium cohort, as parental genotype data were not assessed. SNPs were excluded when: 1) MAF < 0.01; 2) HWE *p*-value < 1×10^-5^; 3) call rate < 0.98; 4) SNPs had duplicate positions; 5) SNPs had 3 or more alleles; and 6) non-ACGT SNPs on the autosomes. The non-transmitted allele status was defined by generating a single transmission-disequilibrium test (TDT) pseudo-control genotype for each child (given the two parents; Plink– tucc option) after defining all children as being cases (Clayton, 1999). To determine the maternal and paternal transmission of haplotypes, the transmitted and non-transmitted alleles datasets were phased (SHAPEITv2.r904). The resulting haplotypes were converted into mother and father non-transmitted homozygous haploid genotypes for polygenic score analyses of non-transmitted alleles per parent.

In an earlier study, we applied integrative multi-omics models to childhood aggression (Hagenbeek, et al., 2022). There, we included transmitted and non-transmitted polygenic scores (PGSs) for childhood aggression and 14 traits with significant (*p* < 0.02) large genetic correlations (≤ -0.40 or ≥ 0.40) with aggression (Ip et al., 2021). As childhood aggression (Ip et al., 2021) had a genetic correlation with ADHD of 1.0028 (se = 0.0732, *p* = 9.39×10^-43^), we included PGSs for the same traits in the current study. PGSs were calculated for transmitted and non-transmitted alleles based on 15 discovery GWA meta-analyses, which omitted NTR and LUMC-Curium from the discovery meta-analysis. The 15 traits comprised childhood aggression (Ip et al., 2021), ADHD (Demontis et al., 2019), Major Depressive Disorder (Wray et al., 2018), Autism Spectrum Disorder (Grove et al., 2019), Loneliness (http://www.nealelab.is/uk-biobank/), Insomnia (Jansen et al., 2019), Self-reported health (http://www.nealelab.is/uk-biobank/), Smoking initiation (ever/never smoked; M. Liu et al., 2019), age of smoking initiation (Watanabe et al., 2019), number of cigarettes per day (M. Liu et al., 2019), Childhood IQ (Benyamin et al., 2014), Educational Attainment (Lee et al., 2018), Age at first Birth (Barban et al., 2016), Wellbeing spectrum (Baselmans et al., 2019), and Intelligence (Savage et al., 2018; also see Hagenbeek, et al., 2022). We calculated the PGSs in LDpred (Vilhjalmsson et al., 2015) which infers the posterior mean effect size of each marker by using a prior on effect sizes and linkage disequilibrium (LD) information from an external reference panel. In 2,500 second-degree unrelated individuals randomly selected from the NTR (the reference panel) the LD weighted betas for all traits were estimated (LDpred v0.9, fraction *F* of variants with non-zero effect *F* = 0.50). For all phenotypes we obtained a transmitted and two non-transmitted (1 non-transmitted for the father and 1 for the mother) PGSs by scoring the LD corrected betas using PLINKv1.9. Thus, in total, 45 PGS were specified for each child. The effects of sex, age at biological sample collection, genotype platform, and the first 10 genetic principal components (PCs) were regressed on the standardized (mean of 0 and standard deviation of 1) PGSs. The residuals of the above regressions were included in the analyses.

### DNA methylation

Genome-wide DNA methylation for the NTR and LUMC-Curium cohorts were measured, according to the manufacturer’s specification, on the Infinium MethylationEPIC BeadChip Kit (Illumina, San Diego, CA, USA; Moran et al., 2016) by the Human Genotyping Facility (HugeF) of ErasmusMC (the Netherlands; http://www.glimdna.org/), using the ZymoResearch EZ DNA Methylation kit (Zymo Research Corp, Irvine, CA, USA) for bisulfite treatment of 500 ng of genomic DNA obtained from buccal swabs. Quality Control (QC) and normalization of the methylation data were carried out with pipelines developed by the Biobank-based Integrative Omics Study (BIOS) consortium as was previously described (Sinke et al., 2019). In short, samples were removed if they failed to pass all five quality criteria of MethylAid (van Iterson et al., 2014) if samples had incorrect relationships (omicsPrint; van Iterson et al., 2018), or sex mismatches (DNAmArray; Min et al., 2018). Methylation probes were set to missing in a sample if they had an intensity value of zero, a bead count < 3, or a detection *p*-value > 0.01. DNA methylation probes were excluded if they overlapped with a SNP or Insertion/Deletion (INDEL), mapped to multiple locations in the genome, or had a success rate < 0.95 across all samples. After QC, 787,711 out of 865,859 sites were retained for analysis for 1424 samples, and cellular proportions were predicted in epithelial tissues using the cell-type deconvolution algorithm Hierarchical Epigenetic Dissection of Intra-Sample-Heterogeneity (HepiDISH) with the reduced partial correlation (RPC; Zheng et al., 2018). Missing methylation β-values were imputed (missMDA;Sinke et al., 2019), and two duplicate samples of NTR participants were excluded. One sample was excluded from the LUMC-Curium dataset because this participant and co-twin were also included in the NTR. The effects of sex, age, percentages of epithelial and natural killer cells, EPIC array row, and bisulfite sample plate were regressed from the methylation β-values of the top 10% most variable methylation sites (78,772 in total) that survived QC (see Hagenbeek, et al., 2022). The resulting residual methylation levels were included in the analyses.

### Metabolomics

Urinary metabolomics data were generated by the Metabolomics Facility of Leiden University (Leiden, the Netherlands) on three platforms: 1) a liquid chromatography-mass spectrometry (LC-MS) platform targeting amines; 2) an LC-MS platform targeting steroid hormones; and 3) a gas chromatography-mass spectrometry (GC-MS) platform targeting organic acids. Subjects were randomized across batches, whilst retaining twin pairs on the same plate. Each batch included a calibration line, QC samples (pooled aliquots from all urine samples; every 10 samples), sample replicates, and blanks. In-house developed algorithms were applied, using the pooled QC samples, to compensate for shifts in the sensitivity of the mass spectrometer across batches. Metabolites were reported as ‘relative response ratios’ (target area/area of internal standard) after QC correction, and metabolites with a relative standard deviation of the QC samples larger than 15% were excluded (6 amines, 3 steroids, and 1 organic acid). Metabolite measurements that fell below the limit of detection/quantification were imputed with half of the value of this limit, or when this limit was unknown with half of the lowest observed level for this metabolite. The effects of sex and age were then regressed from the sample-median normalized and inverse normal rank transformed urinary metabolites.

The metabolomics data (Hagenbeek, et al., 2022) were assessed by ultra-performance liquid chromatography-mass spectrometry (UPLC-MS) for 66 amines and 13 steroids (pre-QC). To measure amines, methanol was added to 5 μL of spiked (with internal standards) urine for protein precipitation and centrifugation of the supernatant. Next, the sample was reconstituted in a borate buffer (pH 8.5) with AQC reagent after sample evaporation (speedvac). Chromatographic separation was achieved by an Agilent 1290 Infinity II LC system (1290 Multicolumn Thermostat and 1290 High-Speed Pump; Agilent Technologies, Waldbronn, Germany) with an Accq-Tag Ultra column (Waters Chromatography B.V., Etten-Leur, The Netherlands). The UPLC was coupled to electrospray ionization on an AB SCIEX quadrupole-ion trap (QTRAP; AB Sciex, Massachusetts, USA). Analytes were monitored in Multiple Reaction Monitoring (MRM) by nominal mass resolution and detected in positive ion mode.

To measure the steroid metabolites, internal standards were added to 90 μL of urine, and samples were filtered with a 0.2μm PTFE membrane. Chromatographic separation was achieved by UPLC (Agilent 1290, San Jose, CA, USA), using an Acquity UPLC CSH C18 column (Waters), with a flow of 0.4 mL/min over a 15 min gradient. Samples were then transferred to a triple quadrupole mass spectrometer (Agilent 6460, San Jose, CA, USA) with electrospray ionization. By switching, positive and negative ion mode analytes were detected in MRM using nominal mass resolution.

Gas chromatography-mass spectrometry (GC-MS) was applied to measure the organic acids. Liquid-liquid extraction with ethyl acetate was applied twice to 50 μL of spiked (with internal standards) urine to extract the organic acids and remove urea. Online derivatization procedures were performed in two steps: 1) oxidation with methoxamine hydrochloride (MeOX, 15 mg/mL in pyridine); and 2) N-Methyl-N-(trimethylsilyl)-trifluoroacetamide (MSTFA) silylation. Chromatographic separation was performed on a 25m (HP-5MS UI) film thickness 30 x 0.25m ID column, with helium as a carrier gas (1,7 mL/min). The mass spectrometer (Agilent Technologies, Waldbronn, Germany), using a single quadrupole with electron impact ionization (70 eV) was operated in SCAN mode (mass range 50–500), using 1 μL of the sample. In total, 21 organic acids were successfully measured.

### Statistical analyses

To examine molecular variation associated with ADHD for PGSs, DNA methylation, and metabolomics, and to build a predictive model for ADHD, we applied an integrative multi-omics method called sparse multi-block supervised analysis (Rohart et al., 2017; Singh et al., 2019). All analyses were performed in R (version 4.1.1), mainly in the mixOmics package (version 6.16.3; Rohart et al., 2017). The analytical design consisted of three steps (Hagenbeek, et al., 2022): 1) three single-omics analyses; 2) two sets of pairwise cross-omics analyses; and 3) a multi-omics analysis (**Figure 1**). The twin sample was randomly split at twin pair level to create two subgroups with 70% of the data for model training (training data), and 30% of the data for model testing (test data; **Table 1**). The performance of final single- and multi-omics models was explored in a follow-up clinical sample. All single- and multi-omics models employed 5-fold cross-validation (CV) with 50 repeats to determine the optimal number of components for each model (the mixOmics perf function), for variable selection in each model (the mixOmics perf function), and for assessment of final model performance in the training data (the mixOmics perf function). For these multi-omics analyses there is a strong requirement of no missing data at any omics level. To keep the input data the same for the single- and multi-omics models, we included participants with complete omics data in all analyses.

**Figure 1.**
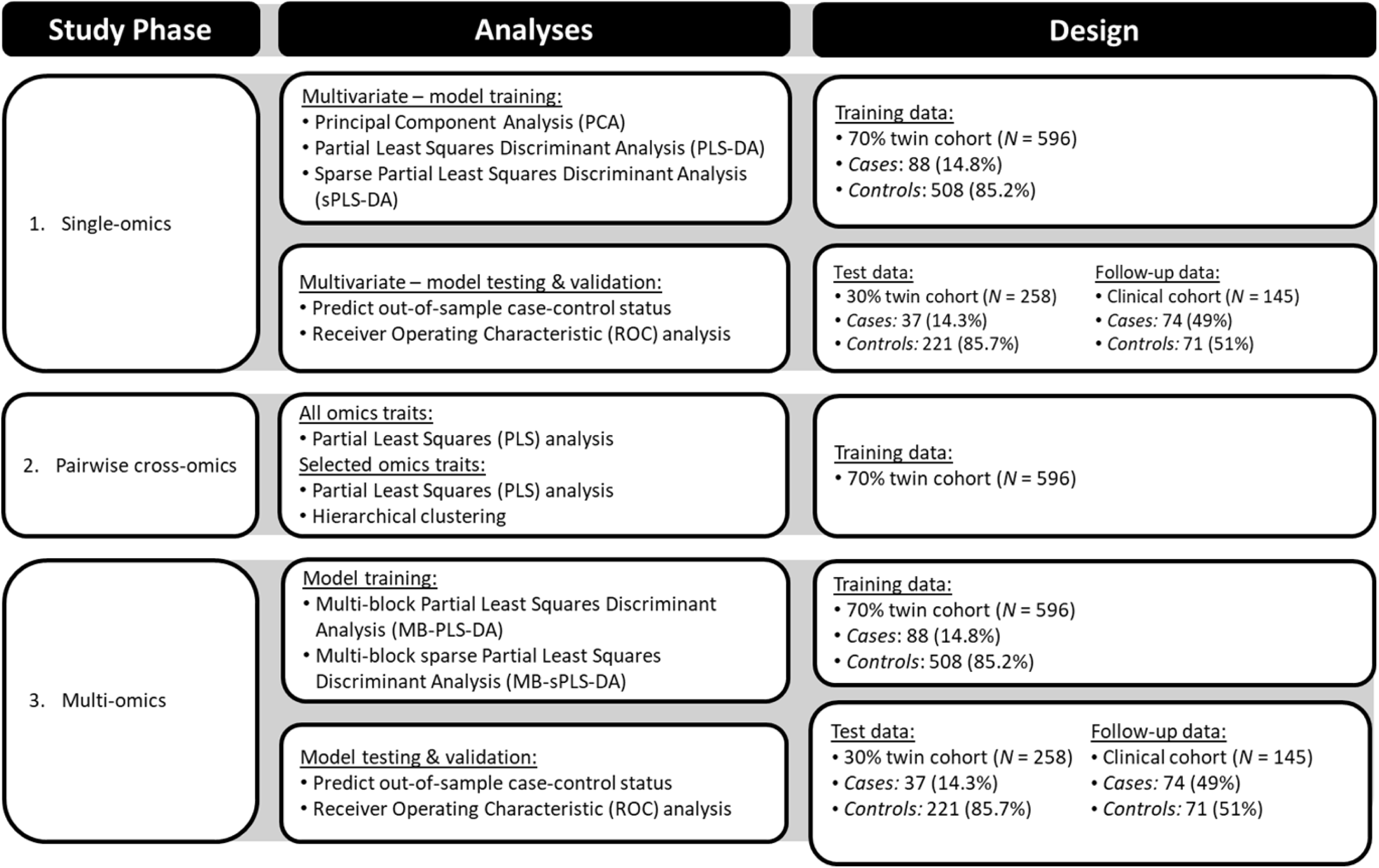
Workflow of the analyses performed in this study. We employed an analytical design consisting of three phases: 1) single-omics analyses; 2) pairwise cross-omics analyses; and 3) multi-omics analyses. First, we built single-omics biomarker panels in the twin cohort, with 70% of the twin data for model training (training data), 30% of the twin data for model testing (test data), and the clinical cohort (follow-up data). Second, the overall pairwise cross-omics correlations among the loading scores of the PLS components for all omics traits and the selected omics variables in the single-omics models were examined in the training data. Third, using the same data split for model training, testing and follow-up, we built multi-omics biomarker panels and described the multi-omics connections of the selected omics traits. The details about the analyses are provided in the **Methods** section.

**Table 1.** Demographics of the training, test, and follow-up data.

|  | Training data (70% twin cohort) |  |  | Test data (30% twin cohort) |  |  | Follow-up data (Clinical cohort) |  |  |
| --- | --- | --- | --- | --- | --- | --- | --- | --- | --- |
|  | Controls | Cases | Total | Controls | Cases | Total | Controls | Cases | Total |
| *N* (%) | 508 (85.2%) | 88<br>(14.8%) | 596<br>(100%) | 221<br>(85.7%) | 37<br>(14.3%) | 258<br>(100%) | 71<br>(49.0%) | 74<br>(51.0%) | 145<br>(100%) |
| *N* (%) complete twin pairs | 227 | 24 | 251 | 98 | 10 | 108 | - | - | - |
| Mean (*SD*) age | 9.5 (1.9) | 9.1 (1.6) | 9.5 (1.9) | 9.6 (2.0) | 9.5 (1.9) | 9.6 (1.9) | 10.5 (1.7) | 9.8 (1.7) | 10.1<br>(1.7) |
| Range age | 5.6 - 12.9 | 5.8 - 12.5 | 5.6 -<br>12.9 | 5.7 - 12.7 | 6.0 - 12.2 | 5.7 -<br>12.7 | 7.0 - 13.3 | 6.3 -<br>13.4 | 6.3 -<br>13.4 |
| *N* (%) females | 257 (50.6%) | 37 (42%) | 294<br>(50.7%) | 113<br>(51.1%) | 14<br>(37.8%) | 127<br>(49.2%) | 18<br>(25.4%) | 19<br>(25.7%) | 37<br>(25.5%) |
| *N* (%) MZ twins | 436 (85.8%) | 65<br>(73.9%) | 501<br>(84.1%) | 175<br>(79.2%) | 30<br>(81.1%) | 205<br>(79.5%) | - | - | - |
| Mean (*SD*) ADHD score<sup>1</sup> | 1.4 (1.6) | 7.1 (1.5) | 2.3 (2.6) | 1.7 (1.6) | 7.2 (1.5) | 2.5 (2.5) | 5.3 (2.5) | 11.9<br>(1.5) | 8.7 (3.9) |

Notes: MZ, monozygotic.
<sup>1</sup> Measured with the mother-rated Attention Deficit/Hyperactivity Problems DSM-oriented scale of the Achenbach System of Empirically Based Assessment (ASEBA) Child Behavior Checklist (CBCL). The ASEBA CBCL Attention Deficit/Hyperactivity Problems DSM-oriented scale scores in the clinical cohort include 90% mother report and 10% father report.

### Step 1: single-omics analyses

Principal Component Analysis (PCA) was run separately for a total of 45 transmitted and maternal and paternal non-transmitted PGSs, 78,772 CpGs in the DNA methylation data, and all 90 metabolites in the training data to obtain a first insight into the dimensionality in each omics level (**Supplementary Figure 1**). Next, Partial Least Square Discriminant Analyses (PLS-DA) and sparse PLS-DA (sPLS-DA) were applied to the training data to assess the ability of each of the three omics levels to correctly classify ADHD status. Cross-validation determined the optimal number of components to include in each PLS-DA model (**Supplementary Table 1**; **Supplementary Figure 2**), determined the optimal number of components, and reduced the number of variables in each omics layer that contribute to each component in the sPLS-DA models (**Supplementary Table 1**; **Supplementary Figure 3**), and assessed the performance of the final sPLS-DA model (**Supplementary Table 1**; **Supplementary Figure 4**). Three types of distance measurements were assessed; 1) centroid distance; 2) Mahalanobis distance and 3) maximum distance. The distance that showed the best predictive performance in the training data was selected for the final models.

We evaluated the out-of-sample predictions in the test and follow-up data (mixOmics predict function) based on the best-performing model (**Supplementary Table 1**). We applied three methods to evaluate how well the final models classified case-control status. First, we employed a balanced misclassification rate, the balanced error rate (BER; false-negative + false-positive rate), that corrects for imbalances in the number of cases and controls. Second, we calculated the sensitivity, specificity, and accuracy of the single-omics models by comparing the predicted cases and controls with the true cases and controls. Third, we assessed the Area Under the Curve (AUC) for each component in both the test and follow-up data with Receiver Operating Characteristic (ROC) analysis.

### Step 2: Pairwise cross-omics analyses

To investigate pairwise cross-omics connections (i.e., PGSs-DNA methylation, PGSs-metabolomics, and DNA methylation-metabolomics) we constructed Partial Least Squares (PLS) regression models in canonical mode in the training data. We calculated the correlations among the loading scores of the PLS components for the selected omics traits. We included 2 components for the PGS-DNA methylation model, 2 components for the PGS-metabolomics model, and 5 components for the DNA methylation-metabolomics model. In total, we ran two sets of PLS models: three models for each pairwise combination of omics data that included all 45 PGSs, 78,772 CpGs, or 90 metabolites. These provide insight into the correlations of the loading scores among all omics traits. The next three models for each pairwise combination of omics data included only the 17 PGSs, 486 CpGs, and 90 metabolites that were selected by the single-omics sPLS-DA models (**Supplementary Data 1**). This second set of models provides insight into the correlations of the loading scores among the omics traits that best contribute to the classification of ADHD cases and controls in the single-omics sPLS-DA models. We performed hierarchical clustering to identify biologically relevant clusters using the Ward linkage algorithm on Euclidean distances of the PLS variates of the PLS models that included the omics traits as selected by the single-omics sPLS-DA models and extracted the two largest clusters for both of the omics levels included in the PLS models (‘dendextend’ R-package; Galili, 2015).

### Step 3: Multi-omics analyses

The multi-omics analysis was conducted through multi-block sPLS-DA (MB-sPLS-DA) in the training data including an empirical design matrix, based on the correlations among the loading scores from the pairwise cross-omics PLS models including all omics traits. The design matrix comprised of correlations of 0.25 between the PGSs and DNA methylation, 0.29 between the PGSs and the metabolites, and 0.23 between DNA methylation and the metabolites (based on Pairwise cross-omics analyses). Cross-validation determined the optimal number of components to include in the MB-PLS-DA models, the number of traits to include per component per omics layer, and the performance of the final MB-sPLS-DA model (**Supplementary Table 2**; **Supplementary Figure 5-7**). The accuracy of out-of-sample case-control status prediction was again evaluated in the test and follow-up data, with final multi-omics models evaluated by their BER and AUCs (ROC curves were calculated per component for each omics layer), and the model sensitivity, specificity, and accuracy were calculated from the confusion matrices.

### Biological characterization

We describe the correlations among the loading scores of the PLS components of the PGSs, CpGs, and metabolites that were selected by the single-omics sPLS-DA and the multi-omics MB-sPLS-DA models to facilitate high correlation patterns suitable for biological interpretation. We performed enrichment analysis for the CpGs selected across our analyses for all phenotypes (618) in the EWAS atlas on the 10^th^ of December 2021 (Xiong et al., 2022). We performed enrichment on 1) the CpGs that were selected by the single-omics sPLS-DA model, 2) the CpGs in the clusters identified by the pairwise cross-omics analyses, 3) the CpGs that were selected for the multi-omics MB-sPLS-DA model, and 4) the CpGs that were included in the high correlation patterns of the multi-omics MB-sPLS-DA. When fewer than 20 CpGs were selected, the trait associations with the CpGs were manually retrieved from the EWAS atlas. We focused the enrichment analysis on the CpGs since there is no similar tool to the EWAS atlas for the metabolites.

## Results

### Single-omics models for ADHD

Based on the sPLS-DA analyses for PGSs, DNA methylation, or metabolomics data we built single-omics prediction models for ADHD case-control status. The single-omics models included 17 PGSs, 486 CpGs, and all 90 metabolites (**Supplementary Table 1**; **Supplementary Data 1**). The 17 selected PGSs in the sPLS-DA model comprised the transmitted PGS for ADHD, smoking initiation, Major Depressive Disorder (MDD), educational attainment (EA), number of cigarettes per day, and insomnia. The non-transmitted maternal PGSs were selected for insomnia, self-reported health, childhood aggression, age of smoking initiation, and childhood IQ. The non-transmitted paternal PGSs were selected for ADHD, intelligence, Autism Spectrum Disorder (ASD), Educational Attainment (EA), number of cigarettes per day, and the age of smoking initiation (**Supplementary Data 1**). Trait enrichment analyses for all 486 selected CpGs in the sPLS-DA model showed the strongest enrichment for glucocorticoid exposure (i.e., based on an EWAS of administration of corticosteroid medication and DNA methylation measured in buccal cells (Braun et al., 2019); N Overlap Differently Methylated CpGs (DMC) = 13, OR = 7.66, *p* = 1.12×10^-14^), and ancestry (DMC = 15, OR = 2.61, *p* = 5.73×10^-6^; **Supplementary Table 3**). Model performance was sub-optimal in the test and follow-up data for all three omics levels (AUC range = [0.45, 0.60], **Supplementary Table 4**).

### PGS-DNA methylation analysis

An average correlation of *r* = 0.23 (*q* = 8.92×10^-16^) was found between the PGSs and DNA-methylation in the loadings scores of the pairwise PLS cross-omics model. For the pairwise model, we applied hierarchical clustering which identified two clusters for the PGS and DNA methylation data (**Figure 2a**; **Supplementary Data 2**). Cluster 1 contains 8 of the PGSs (47.1%) selected by the sPLS-DA model for ADHD, and comprised the transmitted PGSs for ADHD, insomnia, MDD, smoking initiation, and the number of cigarettes per day, the non-transmitted maternal PGSs for insomnia and childhood aggression, and the non-transmitted paternal PGS for ADHD. The 9 PGSs (52.9%) included in cluster 2, comprised the transmitted PGS for EA, the non-transmitted maternal PGSs for age at smoking initiation, childhood IQ, and self-reported health, and the non-transmitted paternal PGSs for age at smoking initiation, ASD, the number of cigarettes per day, and intelligence. The DNA methylation cluster 1 contains 226 (46.5%) of the CpGs selected by the sPLS-DA models for ADHD and showed the strongest trait enrichments for glucocorticoid exposure (DMC = 13, OR = 12.94, *p* = 6.64 ×10^-31^), and trihalomethanes (THM) exposure (DMC = 2, OR = 33.89, *p* = 1.85×10^-11^; **Supplementary Table 5**). The 260 (53.5%) CpGs included in cluster 2 showed the strongest trait enrichment for ancestry (DMC = 13, OR = 4.32, *p* = 3.30×10^-9^), and childhood stress (DMC = 3, OR = 20.69, *p* = 7.20×10^-7^). The correlations of PGSs in clusters 1 and 2 with the CpGs ranged from -0.15 to 0.13 and from -0.20 to 0.21, respectively, and the correlations of the CpGs in clusters 1 and 2 with the PGSs ranged from -0.20 to 0.21 and from -0.17 to 0.16, respectively (**Figure 2a; Supplementary Data 2-3**).

**Figure 2.**
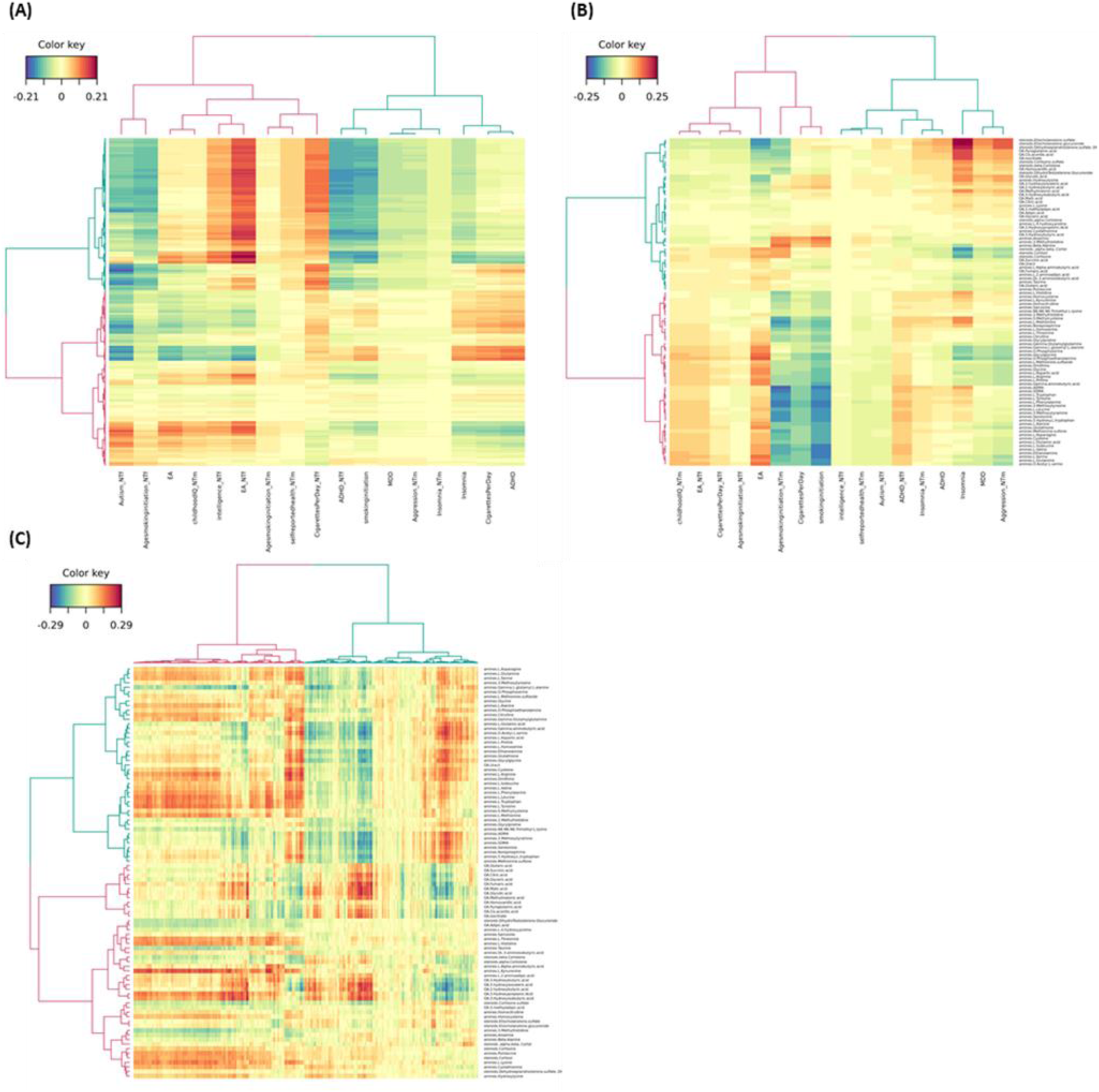
Clustered heatmaps of the correlations among the loading scores of the Partial Least Squares (PLS) components including only the omics traits as selected by the single-omics sparse Partial Least Squares Discriminant Analyses (sPLS-DA). The hierarchical clustering was generated using the Ward linkage algorithm on Euclidean distances of the PLS variates. Two clusters have been identified for each dendrogram (cluster 1 = pink, cluster 2 = blue). Positive correlations among the omics traits have been depicted in red and negative correlation in blue. For the polygenic scores (PGSs), the ‘_NTm’ suffix denotes PGSs non-transmitted by mother, the ‘_NTf’ suffix denotes the PGSs non-transmitted by father, and childhood aggression is abbreviated as “aggression”, Attention-Deficit Hyperactivity Disorder as “ADHD”, Major Depressive Disorder as “MDD”, Autism Spectrum Disorder as “Autism”, and Educational Attainment as “EA”. For the metabolites, the ‘amines.’ prefix indicates these metabolites were measured on the liquid chromatography-mass spectrometry (LC-MS) amines platform, the ‘steroids.’ prefix indicates these metabolites were measured on the LC-MS steroids platform, and the ‘OA.’ prefix indicates these metabolites were measured on the gas chromatography-mass spectrometry (GC-MS) organic acids platform. **(a)** Correlation among the 17 PGSs and 486 CpGs included in the 2-component PGS-DNA methylation PLS model, where the selected CpGs are represented in the rows and the selected PGSs in the columns. The cluster assignments and the full correlation matrix are included in **Supplementary Data 2-3**. (b) Correlations among the 17 PGSs and 90 metabolites included in the 2-component PGS-metabolomics PLS model, where the selected metabolomics traits are represented in the rows and the selected PGSs in the columns. The cluster assignments and the full correlation matrix are included in **Supplementary Data 2** and **Data 4**, respectively. (c) Correlation among the 486 CpGs and 90 metabolites included in the 5-component DNA methylation-metabolomics PLS model, where the selected metabolomics traits are represented in the rows and the selected CpGs in the columns. The cluster assignments and the full correlation matrix are included in **Supplementary Data 2** and **Data 5**.

### PGSs-Metabolomics analysis

Hierarchical cluster of the pairwise PGSs-metabolomics model, with an average correlation between the loading scores of *r* = 0.26 (*q* = 1.02×10^-19^), identified two clusters of PGSs and two clusters of metabolites (**Figure 2b**; **Supplementary Data 2**). The PGS cluster 1 contains 9 of the PGSs (52.9%) selected by the sPLS-DA model for ADHD, comprising the transmitted PGS for ADHD, insomnia, and MDD, the non-transmitted maternal PGSs for insomnia, childhood aggression, and self-reported health, and the non-transmitted paternal PGSs for ADHD, ASD, and intelligence. The 8 PGSs (47.1%) included in cluster 2 comprises the transmitted PGSs for the number of cigarettes per day, EA, smoking initiation, the non-transmitted maternal PGSs for the number of cigarettes per day, age at smoking initiation, and childhood IQ, and the non-transmitted paternal PGSs for EA, and age at smoking initiation. Metabolite cluster 1 contains 48 metabolites (53.3%), all of which are amines (80% of all amines). The remaining 12 amines, as well as all steroids and organic acids, have been included in metabolite cluster 2 (N = 42, 46.7%). Overall, we observed correlations ranging from -0.17 to 0.25 for the PGSs in cluster 1 with metabolites, and from -0.22 to 0.16 for the cluster 2 PGSs with metabolites (**Figure 2b**; **Supplementary Data 2** and **Data 4**). For the metabolites in cluster 1 the correlation ranged between -0.22 to 0.16 with PGS, and for cluster 2 between -0.17 to 0.25.

### DNA methylation-Metabolomics analysis

The pairwise cross-omics DNA methylation-metabolomics model showed an average correlation of 0.23 (*q* = 3.95×10^-37^) and hierarchical clustering identified two clusters of CpGs and of metabolites (**Figure 2c**; **Supplementary Data 2**). The DNA methylation cluster 1 contains 241 (49.6%) of the CpGs selected by the sPLS-DA models for ADHD, and the strongest trait enrichments for these CpGs were observed for glucocorticoid exposure (DMC = 13, OR = 12.08, *p* = 7.63×10^-30^), and THM exposure (DMC = 2, OR =31.79, *p* = 3.22×10^-11^; **Supplementary Table 5**). The 245 (50.4%) CpGs included in cluster 2 show the strongest trait enrichment for ancestry (DMC = 13, OR = 4.60, *p* = 9.96×10^-10^), childhood stress (OR = 21.97, *p* = 5.12×10^-7^), and household socioeconomic status in childhood (DMC= 3, OR = 18.34, *p* = 7.20×10^-7^). Metabolite cluster 1 contains 43 (47.8%) of the metabolites, including 42 amines and 1 organic acid, while metabolite cluster 2 contains 47 metabolites (52.2%), including 18 amines, 19 organic acids, and all 10 steroids. The correlation of the CpGs included in cluster 1 with metabolites range from -0.23 to 0.29 (**Figure 2c**; **Supplementary Data 2** and **Data 5**). The CpGs included in cluster 2 have correlations ranging from -0.27 to 0.28 with metabolites. For the metabolites in cluster 1, the correlations with CpGs ranged from -0.26 to 0.25, and for cluster 2 from -0.27 to 0.29.

### Multi-omics model for ADHD

We built a multi-omics panel for ADHD based on a multi-block sPLS-DA (MB-sPLS-DA) model including PGSs, DNA methylation, and metabolomics data. The 4-component model included in total 30 PGSs, 143 CpGs, and all 90 metabolites (**Supplementary Table 2**; **Supplementary Data 6**). The number of components and number of variables per omics layer were determined by the single-omics models. The included variables in the multi-omics model were selected based on CV. The multi-omics model comprised the transmitted PGS for ADHD, aggression, cigarettes per day, insomnia, loneliness, childhood IQ, self-reported health, age at first birth, intelligence, smoking initiation, EA, MDD, and wellbeing. The non-transmitted maternal PGSs were selected for aggression, cigarettes per day, insomnia, loneliness, childhood IQ, self-reported health, and age smoking initiation. The non-transmitted paternal PGSs were selected for ADHD, aggression, cigarettes per day, insomnia, loneliness, age at first birth, intelligence, smoking initiation, age smoking initiation, and ASD. Trait enrichment analyses for all selected CpGs in the MB-sPLS-DA model showed the strongest enrichment for childhood stress (DMC = 3, OR = 38.00, p = 2.25×10-8), and respiratory allergies (AR; DMC = 2, OR = 28.48, *p* = 1.58×10^-5^; **Supplementary Table 6**). The multi-omics prediction in the test data showed a better prediction (0.42 ≤ BER ≤ 0.51; 0.43 ≤ AUC ≤ 0.62) as compared to single-omics models, while prediction in the follow-up data was similar to those observed for the single-omics models (0.49 ≤ BER ≤ 0.55; 0.50 ≤ AUC ≤ 0.53; **Supplementary Table 7; Supplementary Table 4).**

The average correlations among the loading scores of the PLS components between each omics layer in the multi-omics model were *r* = 0.17 (*q* = 1.41×10^-16^) for PGSs-DNA methylation, *r* = 0.14 (*q* = 1.24×10^-11^) for PGSs-metabolomics, and *r* = 0.20 (*q* = 1.1×10^-21^) for DNA methylation-metabolomics. We observed high absolute correlations (*r* ≥ 0.60) between 4 selected PGSs, 19 CpGs, and 10 metabolites, which can be summarized in three sets of correlational patterns (**Figure 3**; **Supplementary Data 7**; **Supplementary Data 8**). Correlation pattern 1 comprises correlations of CpGs located in the STAP2 gene with the transmitted ADHD PGS (r range: -0.62- -0.61), the transmitted self-reported health PGS (*r* range = [0.63, 0.66], and eleven amino acids (*r* range = [-0.71, -0.60]). Correlation pattern 2 comprises correlations between 8 CpGs and the non-transmitted paternal cigarettes per day PGS (*r* range = [-0.70, -0.61]). Two of these CpGs (cg15914316 and cg25352924) are located in the *MAD1L1* gene (chromosome 7). Correlation pattern 3 comprises correlations between five CpGs and the transmitted childhood IQ PGS (*r* range = [-0.60, 0.67]). Three of these five CpGs are located in genes coding for transmembrane proteins (*C7orf49*, *TMEM100*, *TMEM135*, and *TMEM140*). The other two CpGs (cg10864680 and cg06843189) are located in *TNFSF13B* (chromosome 13) and *NR3C2* (chromosome 4), respectively.

**Figure 3.**
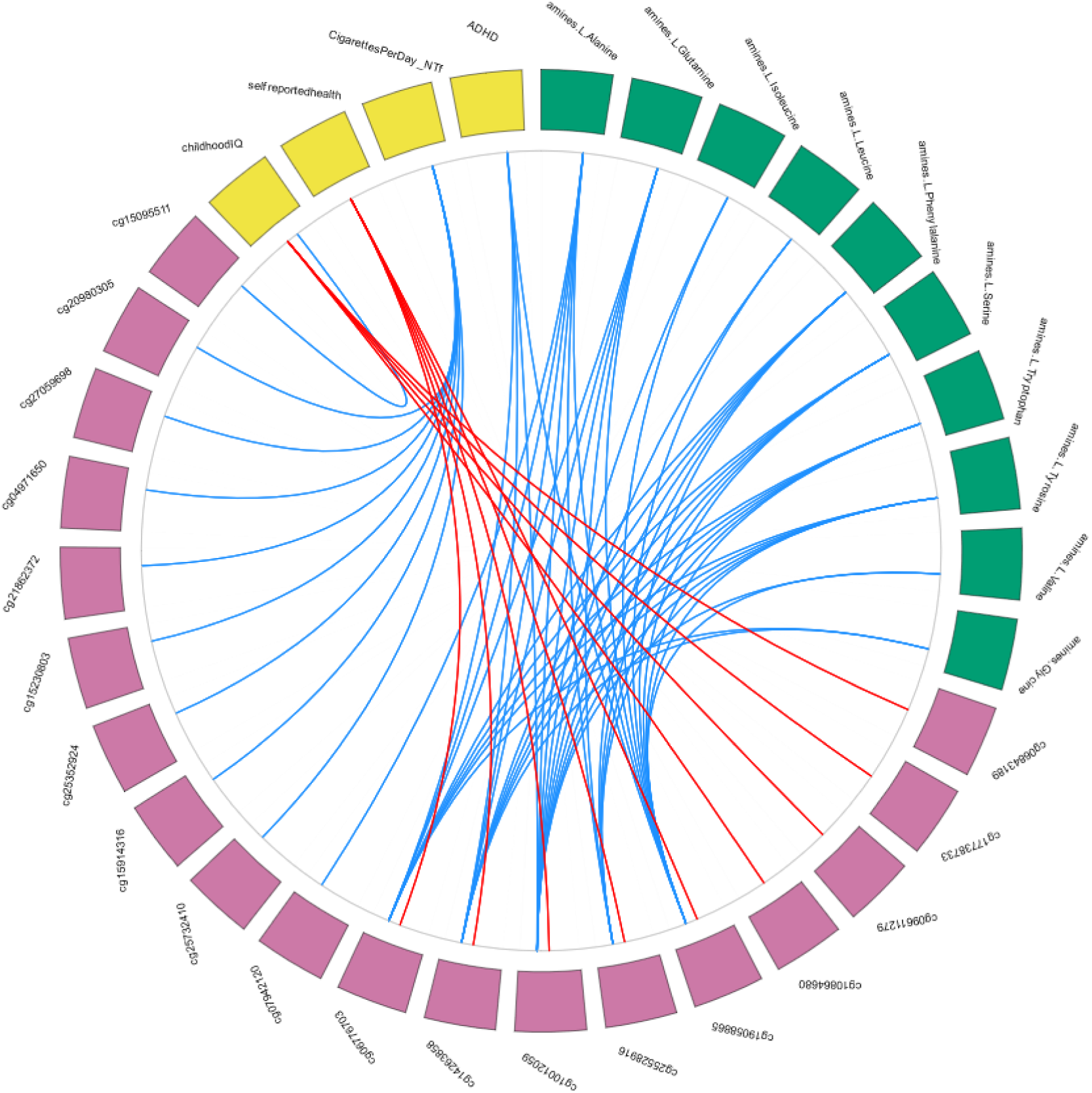
High cross-omics correlations among the loading scores of the PLS components of the multi-omics traits identified in the 4-component multi-block sparse Partial Least Squares Discriminant Analysis (MB-sPLS-DA) model. The outer ring depicts the PGSs, CpGs, and metabolites in yellow, pink, and green, respectively. For the polygenic scores (PGSs), Educational Attainment is abbreviated as “EA”, and the ‘_NTm’ suffix denotes the PGSs non-transmitted by mother. The inner plot depicts the correlations among the omics traits. For the metabolites, the ‘amines.’ prefix indicates these metabolites were measured on the liquid chromatography-mass spectrometry (LC-MS) amines platform, and the ‘OA.’ prefix indicates these metabolites were measured on the gas chromatography-mass spectrometry (GC-MS) organic acids platform. Here, only high absolute correlations (*r* ≥ 0.60) between traits of at least two omics levels are depicted, with blue lines reflecting negative correlations and red lines positive correlations. Correlations are averaged across all components in the MB-sPLS-DA model. The full correlation matrix is included in **Supplementary Data 7** and the correlational patterns are included in **Supplementary Data 8**.

## Discussion

We present an integrative multi-omics analysis of childhood ADHD based on genomics, epigenomics, and metabolomics data to identify multi-omics biomarkers and to investigate the connections among the different omics levels. The single-omics models selected 17 PGSs, 486 CpGs, and 90 metabolites and the multi-omics model selected 30 PGSs, 143 CpGs, and 90 metabolites, and several of the selected variables support known associations with ADHD. The out-of-sample predictions were sub-optimal in both the single- and multi-omics models (BER ranges [0.40, 0.57] and [0.42, 0.55], respectively). We evaluate the pairwise variable associations and found that the averaged correlation values range from 0.23 to 0.29. The connections of the ADHD biomarkers in the multi-omics model highlighted several possibly involved genomic regions and further supported previously identified markers such as the gene *MAD1L1,* childhood stress, and glucocorticoid exposure.

The single-omics models selected 17 PGS based on 12 phenotypes, while the multi-omics model selected 30 PGSs comprising all 15 phenotypes. Transmitted PGSs were selected for ADHD, insomnia, MDD, EA, smoking initiation, and cigarettes per day. These traits are genetically correlated with ADHD (Demontis et al., 2019, 2021). One study that looked at the intergenerational transmission of EA and ADHD found that transmitted PGSs influenced ADHD behavior at school (de Zeeuw et al., 2020). For traits such as aggression, self-reported health, age of smoking initiation, intelligence, childhood IQ, and autism only non-transmitted PGSs were selected. This may represent an important observation: the genotype of the parents can help predict the ADHD status of their offspring when we consider their non-transmitted scores across a broad range of traits. We note that the predictive accuracy of the model is currently low but may improve when PGSs improve with increasing GWAS sample size.

The CpGs in the single-omics model were enriched for multiple traits and disorders previously associated with ADHD, such as glucocorticoid exposure, Klinefelter syndrome, and childhood stress (Friedrichs et al., 2012; Grimm et al., 2020; Harpin, 2005; Lo-Castro et al., 2011; Xiong et al., 2022). In the single- and multi-omics models, we observed enrichment for traits involved in diet exposure, such as glucocorticoid and vitamin B12 levels. We also found associations for THM exposure, a class of chemical compounds that can be present when water is disinfected with chlorine (Hood, 2005). Still, we note that the number of overlapping DMCs with all known DMCs in the EWAS atlas was usually relatively small. The multi-omics model selected two novel CpGs located in the gene *MAD1L1* previously linked to ADHD in blood and saliva tissue (Goodman et al., 2020; Mooney et al., 2020; Rovira, Sanchez-Mora, et al., 2020). Another finding for rs11982272 located in *MAD1L1* has previously been reported as a shared risk locus between ADHD and disruptive behavior disorders (DBD) but was only moderately associated with ADHD in a large GWAS (Demontis et al., 2021). *MAD1L1* was also identified in studies focusing on schizophrenia and bipolar disorder, suggesting that *MAD1L1* is a risk gene for several psychiatric disorders (Ikeda et al., 2018; Ripke et al., 2014).

The CpGs in the multi-omics models were connected with the non-transmitted paternal cigarettes per day PGS in our study. The connection suggests that the association between ADHD and the methylation around *MAD1L1* might be caused by the effects of paternal smoking or that paternal smoking is a confounder between ADHD and the methylation around *MAD1L1* (e.g., parental smoking leads to both ADHD and DNA methylation, but there is no connection between the DNA methylation and ADHD). Still, to our knowledge this is the first observation of paternal smoking effects on ADHD and paternal smoking should merit investigation in future studies. An earlier multi-omics study identified several genes whose expression or DNA methylation level in the (fetal) brain mediates the effects of genetic variants on ADHD, including a gene for a transmembrane protein *TMEM125* (Hammerschlag et al., 2020). We observed four CpGs located in three genes of this same gene family (*TMEM100, TMEM135*, and *TMEM140*) had a connection with the transmitted childhood IQ PGS. The findings of *MAD1L1* and the *TMEM* family support the results of the previous studies and imply an effect of these genomics regions on ADHD.

Both the single- and multi-omics models selected all 90 metabolites, which is not unexpected since the data were generated on targeted platforms, covering metabolites previously associated with aggression (Hagenbeek et al., 2016), and aggression is the trait which is one of the strongest associated variables with ADHD in childhood (Bartels et al., 2018). Each metabolite by itself has a small effect and single and multi-omics models are not equipped to identify these small effects, but a combination of all metabolites discriminates better between ADHD cases and controls.

We observed three connection patterns in our multi-omics analysis where the omics variables showed correlation coefficients of *r* ≥ 0.60. As a result, we can make several observations about these patterns. First, only amines (mostly essential amino acids) showed high negative correlations with CpGs (*r* range = [-0.71, -0.60]) and we observed no high correlations between the organic acids and the steroids, and the other omics levels. Recent studies have linked several of these essential amino acids, such as glycine, serine, leucine, and valine, to childhood ADHD (Anand et al., 2021; Yu et al., 2021). Essential amino acids that come from dietary intake. This suggests a relationship between diet, DNA methylation, and metabolites involved in ADHD and that DNA methylation mediates the effect of diet on metabolite levels. (Dekkers et al., 2016). A previous study suggested *IGF2* methylation mediates the effect of prenatal diet on ADHD symptoms in childhood (Rijlaarsdam et al., 2017). Our observation concerning the amino acids supports hypotheses that essential amino acids affect childhood ADHD, and that diet might help to improve the symptoms of ADHD (Anand et al., 2021; Hamza et al., 2019; Yu et al., 2021). We would like to point out that, even though we highlight these associations, we cannot make claims about cause and effect. Our study identified associations between ADHD and amino acids and which indicate that it is worthwhile to further examine the possibility of causal relations among diet, DNA methylation, and amino acids.

Another multi-omics connection pattern comprised the transmitted PGSs for ADHD and self-reported health with 6 CpGs and 10 amino acids. The 6 CpGs are located in the *STAP2* gene (Signal Transducing Adaptor Family Member 2) on chromosome 19. No SNPs or CpGs in or around this gene were associated with ADHD in GWA or EWA studies, suggesting *STAP2* is a novel genomic region that could be involved in ADHD (Demontis et al., 2019, 2021; Goodman et al., 2020; Mooney et al., 2020; Neumann et al., 2020; Rovira, Demontis, et al., 2020; Rovira, Sanchez-Mora, et al., 2020; Walton, 2019; Walton et al., 2017). The 6 CpGs and the STAP2 gene have been associated with other traits such as smoking behavior, aging, and Type 2 Diabetes in blood (Xiong et al., 2022; **Supplementary Data 8**). Although we observed no connection between these CpGs and the smoking-related PGSs, it should be taken into account that smoking PGSs are an imperfect measure of overall smoking exposure.

## Limitations

A challenge in machine learning via cross validations is the risk of leaky preprocessing: data processing that inadvertently causes information to leak from the training set to the test set (Whalen et al., 2021). Even though our methods do not entirely consist of machine learning, leaky preprocessing could be present. We preselected the top 10% most variable methylation sites before the split in training and test set to limit the computational burden of our method. We selected these methylation sites solely based on the criterion of variability, regardless of any association of these probes with a phenotype. Since we randomly selected test and training set samples, we may assume the same distributions in both sub-samples.

A source of confounding in the ACTION cohort are the effects of puberty. We do not, however, expect puberty to affect the results in any major way, given the ages of the subjects and the fact that only three females had had their first period.

A major limitation of most multi-block multi-omics methods, including MB-sPLS-DA, is that these methods do not allow for any missing values in the training data on any of the included omics levels. For our study, this meant we had to exclude 367 participants (almost one third of the original sample) because of missing or poor quality data in at least one of the omics levels. Allowing missing data is an important improvement that multi-omics methods need to address in the future.

## Strengths

In contrast to previous multi-omics studies for ADHD, which relied on sequential integration of summary statistics (Cabana-Domínguez et al., 2022; Cheng et al., 2020; Hammerschlag et al., 2020), our multi-block integration approach allowed simultaneous modeling of multiple omics levels and gain insights into the relationships among the different omics levels (Wörheide et al., 2021). Unsupervised multi-omics methods, such as PLS structural equation modeling (PLS-SEM; Csala et al., 2020) or Multi-Omics Factor Analysis (MOFA, Argelaguet et al., 2018), constitute powerful alternatives to explore the relationships among omics levels, but are not appropriate for classification of groups, as was one of the aims of this study. Another asset of our study is that by employing an integrative multi-omics design with harmonized sample collection and measurement protocols in the twin and the clinical groups, we reduced the detrimental heterogeneity due to, for example, differences in sample size for different omics layers, or study protocols as far as possible (Wörheide et al., 2021). Problems with design and technical heterogeneity are not uncommon in sequential integration methods. In the current study, such undesirable sources of technical heterogeneity were minimized by our choice of analytical design and by streamlining the collection protocol for biological samples across twins from the NTR and participants from LUMC-Curium. We collected the biological samples in both cohorts in parallel and generated the omics data for both cohorts together. Other than the difference in cohort type, population vs. clinical, the main difference between these cohorts is the absence of genotyping information for the parents of the children included in LUMC-Curium and non-transmitted PGSs could not be assessed in the clinical cohort.

## Conclusion

Our study underscores the complexity of ADHD, as it remains a challenge to detect multi-omics biomarkers and indicates that clinically useful prediction models based on omics data are not viable yet. We showed how different omics levels involved in ADHD are connected to each other, leading to new hypotheses about cross-omics interactions. The connection patterns reproduced previously identified associations with ADHD, including the *MAD1L1* gene and glucocorticoid exposure, while also highlighting new interesting genomic regions such as the *STAP2* gene. Multi-omics methods are expected to become increasingly important in the translation from association studies to clinical applications and it is of value to continue to improve them. The integrated approach we applied in this study requires raw data analysis and demands that all participants are measured across all omics levels. Solving the challenge of missing data in multi-omics approaches will lead to increased sample sizes and increased power in future projects. Multi-omics might be especially promising for complex traits such as ADHD, where clinicians have been struggling to find fitting treatments for patients and struggle to predict the persistency of ADHD (Caye et al., 2019). By improving methods and enhancing models, as well as by including more omics levels such as transcriptomics, proteomics, and exposome (Cheng et al., 2020; J. Liu et al., 2021), the identification of multi-omics biomarkers will eventually aid in diagnosing and improving personalized treatment plans for persons diagnosed with ADHD.

## Ethics statement

Parents provided written informed consent for their children and twin parents provided written informed consent for their own participation. Study approval was obtained from the Central Ethics Committee on Research Involving Human Subjects of the VU University Medical Center, Amsterdam (NTR 25th of May 2007 and ACTION 2013/41 and 2014.252), an Institutional Review Board certified by the U.S. Office of Human Research Protections (IRB number IRB00002991 under Federal-wide Assurance-FWA00017598; IRB/institute codes), and the Medical Ethical Committee of Leiden University Medical Center (B15.017, B17.031, B17.032 and B17.040).

## Author Contributions

Conceptualization, F.A.H., R.P., A.C.H., V.F., R.R.J.M.V., M.B., T.H., D.I.B.; Data curation, R.P., C.E.M.vB., J.J.H., J.vD.; Formal analysis, N.H., F.A.H., R.P., J.J.H.; Funding acquisition, A.C.H., V.F., R.R.J.M.V., M.B., T.H., D.I.B.; Investigation, F.A.H., A.C.H., P.J.R., E.A.E., R.R.J.M.V., M.B., T.H., J.vD., D.I.B.; Methodology, S.D.; Project administration, F.A.H., D.I.B.; Supervision, R.P., J.vD., D.I.B.; Visualization, N.H., F.A.H.; Writing – original draft, N.H., F.A.H., R.P., J.J.H., J.vD., D.I.B.; Writing – review and editing, all authors. All authors have read and agreed to the published version of the manuscript.

## Data Availability

The standardized protocol for large scale collection of urine and buccal-cell samples in the home situation as developed for the ACTION Biomarker Study in children available at http://www.action-euproject.eu/content/data-protocols. The data of the Netherlands Twin Register (NTR) ACTION Biomarker Study may be accessed, upon approval of the data access committee, through the NTR (https://tweelingenregister.vu.nl/information_for_researchers/working-with-ntr-data)

## Acknowledgments

The Netherlands Twin Register (NTR) warmly thanks all twin families for their participation. LUMC-Curium thanks all patients and their parents for participating, and clinicians for their support. NTR and LUMC-Curium are grateful to all researchers involved in the data collection for the ACTION Biomarker Study.

## Data Availability Statement

The standardized protocol for large-scale collection of urine and buccal-cell samples in the home situation as developed for the ACTION Biomarker Study in children is available at http://www.action-euproject.eu/content/data-protocols. The data of the Netherlands Twin Register (NTR) ACTION Biomarker Study may be requested through the NTR (https://tweelingenregister.vu.nl/information_for_researchers/working-with-ntr-data).

