## Supplement Figures and Tables for "Integrative multi-omics analysis of genomic, epigenomic, and metabolomics data leads to new insights for Attention-Deficit/Hyperactivity Disorder"

***Supplementary Material***

### 1. Supplementary Figures

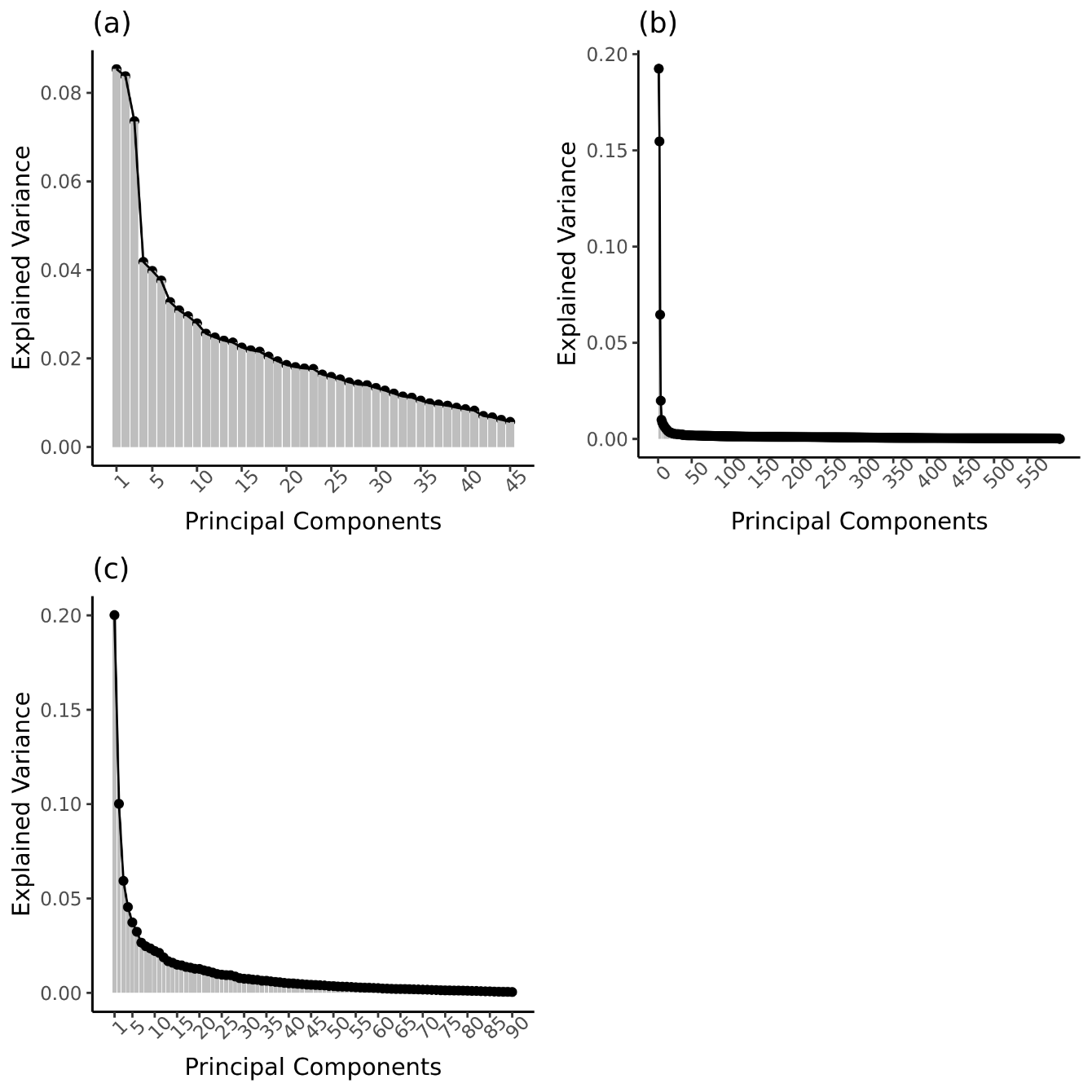

**Supplementary Figure 1.** Scree plots of the Principal Components Analysis (PCA) of the three omics levels. **(a)** Scree plot of the transmitted and non-transmitted polygenic scores (PGSs). **(b)** Scree plot of the DNA methylation data. Here only the 595 Principal Components (PCs) with eigenvalues equal to or larger than one are depicted. **(c)** Scree plot of the metabolomics data.

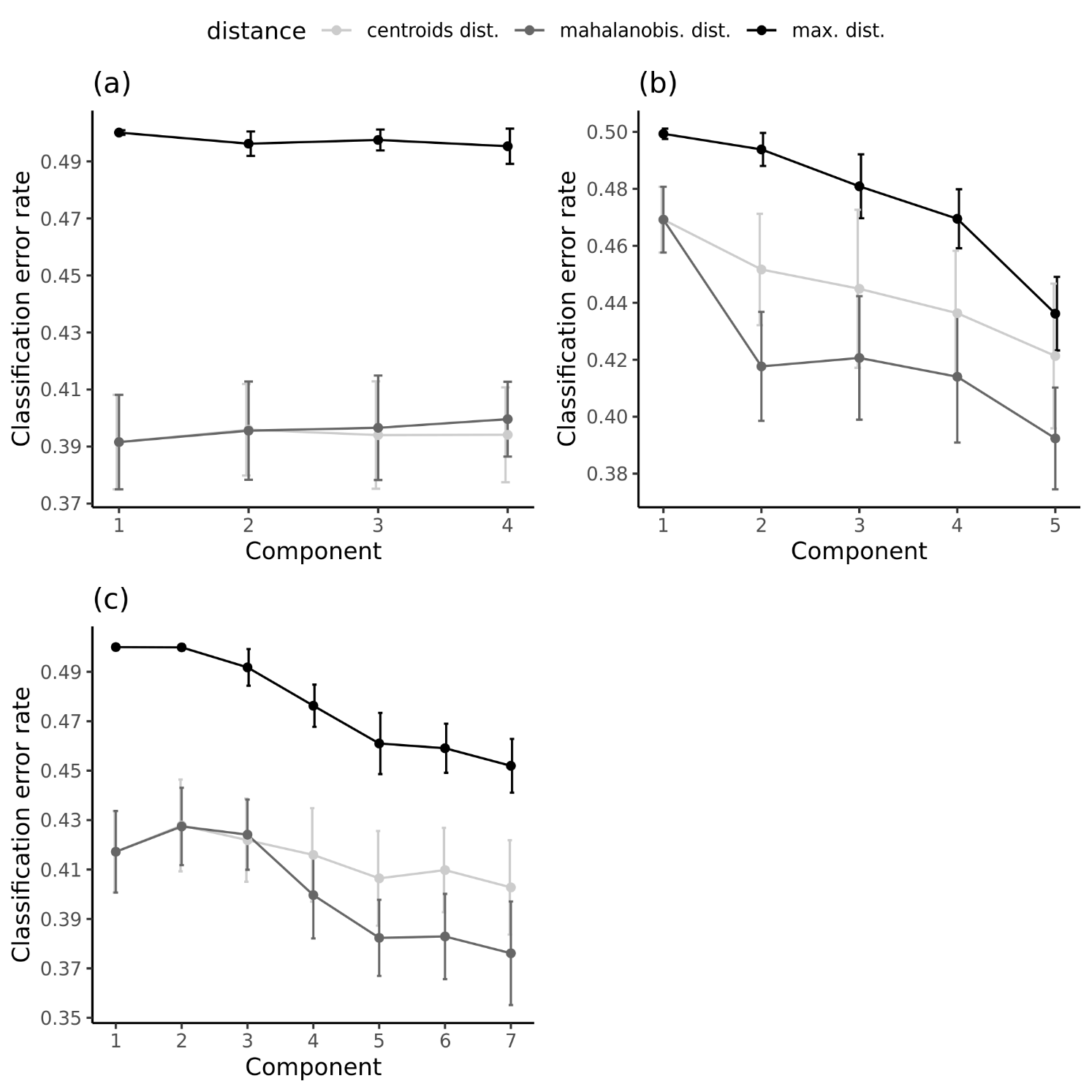

**Supplementary Figure 2.** Balanced error rates (BER) for the number of components to retain in the Partial Least Squares Discriminant Analyses (PLS-DA) to predict ADHD cases and controls in the training data. **(a)** BER for the 4-component PLS-DA model of the transmitted and non-transmitted polygenic scores (PGSs). **(b)** BER for the 5-component PLS-DA model of the DNA methylation data. **(c)** BER for the 7-component PLS-DA model of the metabolomics data. The BER for the centroids distance is depicted in light grey, for the Mahalanobis distance in dark grey, and of the maximum distance in black.

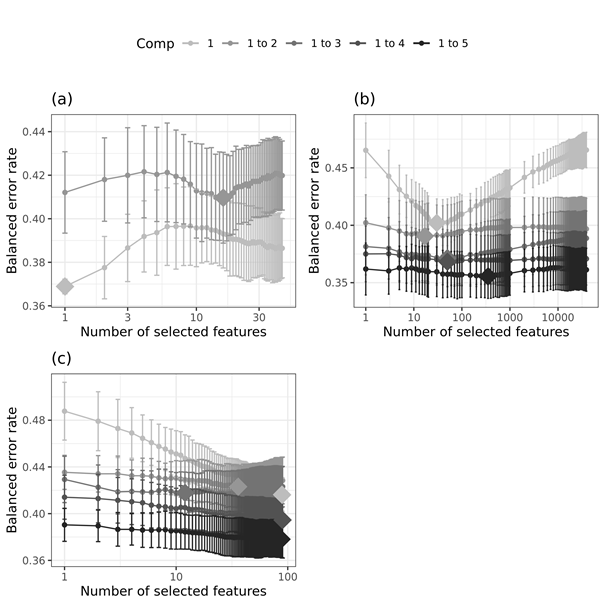

**Supplementary Figure 3.** Balanced error rates (BER) for the number of omics variables per component to retain in the sparse Partial Least Squares Discriminant Analyses (sPLS-DA) models to predict ADHD cases and controls in the training data. **(a)** BER, with centroids prediction distance, of the 2-component sPLS-DA model for the transmitted and non-transmitted polygenic scores (PGSs). **(b)** BER, with Mahalanobis prediction distance, of the 5-component sPLS-DA model for the DNA methylation data. **(c)** BER, with Mahalanobis prediction distance, of the 5-component sPLS-DA model for the metabolomics data. The BERs for each component are given in shades of gray, ranging from the lightest gray for one component to black for five components. The diamonds represent the selected number of features per component by the sPLS-DA model.

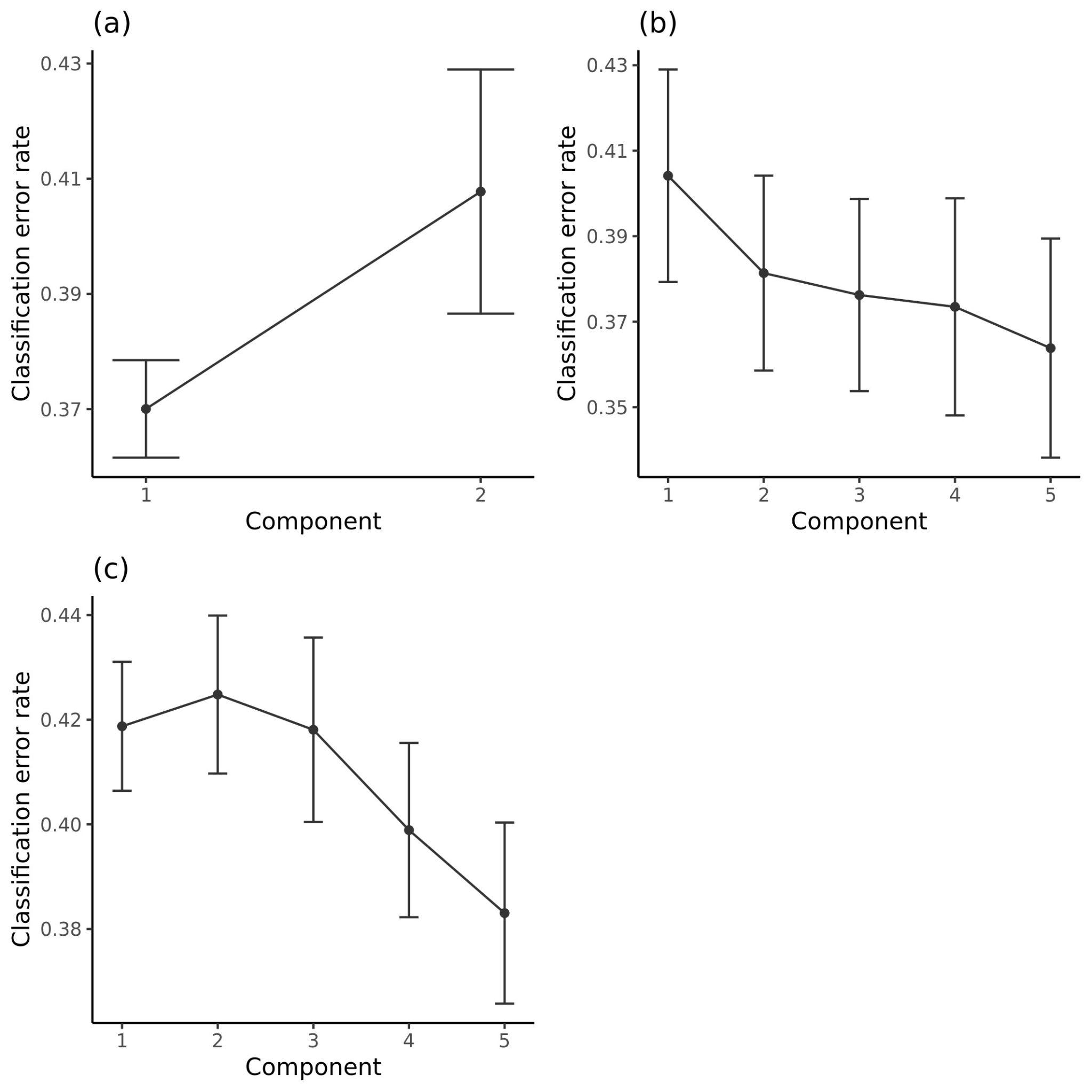

**Supplementary Figure 4.** Balanced error rates (BER) for the number of components to retain in the final sparse Partial Least Squares Discriminant Analyses (sPLS-DA) to predict ADHD cases and controls in the training data. **(a)** BER, with centroids prediction distance, of the 2-component final sPLS-DA model for the transmitted and non-transmitted polygenic scores (PGSs). **(b)** BER, with Mahalanobis prediction distance, of the 5-component final sPLS-DA model for the DNA methylation data. **(c)** BER, with centroids prediction distance, of the 5-component final sPLS-DA model for the metabolomics data.

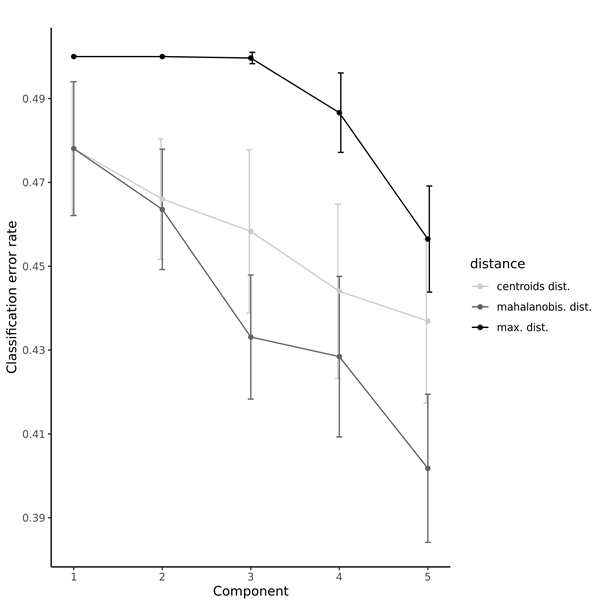

**Supplementary Figure 5.** Balanced error rates (BER), with Mahalanobis prediction distance, for the number of components to retain in the final 5-component multi-block Partial Least Squares Discriminant Analyses (MB-PLS-DA) model to predict ADHD cases and controls in the training data. The BER for the centroids distance is depicted in light grey, for the Mahalanobis distance in dark grey, and of the maximum distance in black.

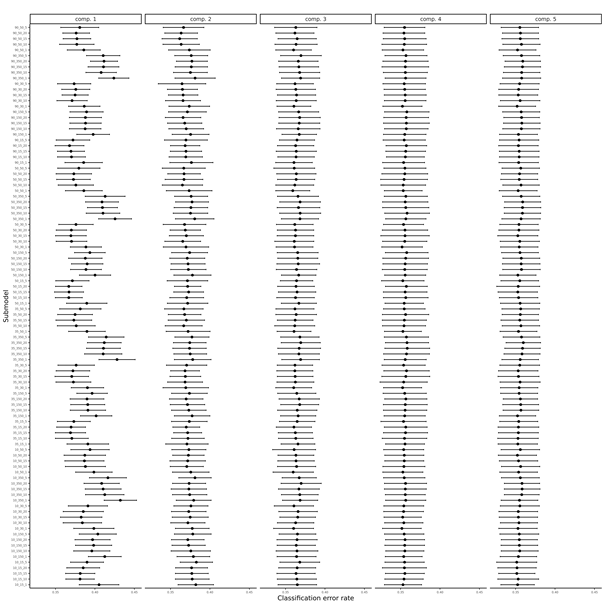

**Supplementary Figure 6.** Balanced error rates (BER) for the number of variables to retain per omics layer and per component in the 5-component multi-block sparse Partial Least Squares Discriminant Analysis (MB-sPLS-DA) model to predict ADHD cases and controls in the training data. In the model tuning 100 possible sub-models were evaluated, each with a different combination of variables to select per omics level.

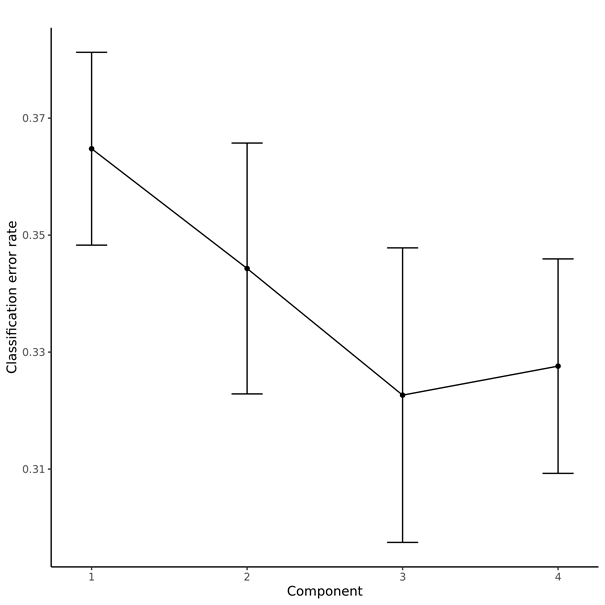

**Supplementary Figure 7.** Balanced error rates (BER) for the number of components to retain in the 4-component multi-block sparse Partial Least Squares Discriminant Analysis (MB-sPLS-DA) model to predict ADHD cases and controls in the training data.

### 2. Supplementary Tables

**Supplementary Table 1.** Overview of the number of components and omics variables per component to retain in the (sparse) Partial Least Squares Discriminant Analyses ((s)PLS-DA) for the three omics levels. For the PLS-DA analyses the number of components used to initialize the models is given, and the optimal number of components (N comp) selected through 5-fold Cross Validation (CV) with 50 repeats. For the tuning of the sPLS-DA models, the number of components and number of omics variables per component to initialize the model tuning is given, followed by the optimal number of omics variables per component and optimal number of components selected through 5-fold CV with 50 repeats. For the final sPLS-DA models the number of components and number of omics variables per component are given, followed by the optimal number of components selected through 5-fold CV with 50 repeats.

|  |  | **PLS-DA** | | **Tune sPLS-DA** | | | | | **Final sPLS-DA** | | |
| --- | --- | --- | --- | --- | --- | --- | --- | --- | --- | --- | --- |
|  | **N variables** | **N comp** | **Optimal N comp** | **N comp** | **Prediction distance** | **N variables per comp** | **Optimal N variables/comp** | **Optimal N comp** | **N comp** | **N variables per comp** | **Optimal N comp** |
| Polygenic scores | 45 | 4 | 1 | 2 | centroids | 1 through 45 | 1, 16 | 2 | 2 | 1, 16 | 2 |
| DNA methylation | 78,772 | 5 | 5 | 5 | mahalanobis | 1, 3, 5, 7, 9, 11, 13, 15, 17, 19, 20, 30, 40, 50, 60, 70, 80, 90, 100, 150, 200, 250, 300, 350, 400, 450, 500, 550, 600, 650, 700, 750, 800, 850, 900, 950, 1000, 2000, 3000, 4000, 5000, 6000, 7000, 8000, 9000, 10000, 11000, 12000, 13000, 14000, 15000, 16000, 17000, 18000, 19000, 20000, 21000, 22000, 23000, 24000, 25000, 26000, 27000, 28000, 29000, 30000, 31000, 32000, 33000, 34000, 35000, 36000, 37000, 38000, 39000 | 30, 17, 50 50, 350 | 5 | 5 | 30, 17, 50 50, 350 | 5 |
| Metabolomics | 90 | 7 | 5 | 5 | mahalanobis | 1 through 90 | 89, 36, 12, 90, 88 | 5 | 5 | 89, 36, 12, 90, 88 | 5 |

**Supplementary Table 2.** Overview of the number of components and number of omics variables per component to retain in the multi-omics (sparse) multi-block Partial Least Squares Discriminant Analyses (MB-(s)PLS-DA). For the MB-PLS-DA analyses the number of components used to initialize the model is given, this is based on the number of components in the final sPLS-DA single-omics models with the largest number of components (i.e., 5 components for the DNA Methylation data; see **Supplementary Table 1**). The optimal number of components (N comp) selected through 5-fold Cross Validation (CV) with 50 repeats is also provided for the MB-PLS-DA analyses. For the tuning of the MB-sPLS-DA models, the number of components and number of omics variables per component to initialize the model tuning is given, followed by the optimal number of omics variables per component and optimal number of components selected through 5-fold CV with 50 repeats. For the final MB-sPLS-DA models the number of components and number of omics variables per component are given, followed by the optimal number of components selected through 5-fold CV with 50 repeats.

| **Data** |  | **MB-PLS-DA** | | **Tune MB-sPLS-DA** | | | | | **Final MB-sPLS-DA** | | |
| --- | --- | --- | --- | --- | --- | --- | --- | --- | --- | --- | --- |
| **Omics layer** | **N variables** | **N comp** | **Optimal N comp** | **N comp** | **Prediction distance** | **N variables/comp** | **Optimal N variables/comp** | **Optimal N comp** | **N comp** | **N variables/comp** | **Optimal N comp** |
| Polygenic scores | 45 | 5 | 5 | 5 | mahalanobis | 1, 5, 10 ,15, 20 | 20, 15, 1, 1 | 4 | 4 | 20, 15, 1, 1 | 4 |
| DNA methylation | 78,772 |  |  |  |  | 15, 30, 50, 150, 350 | 15, 50, 50, 30 |  |  | 15, 50, 50, 30 |  |
| Metabolomics | 90 |  |  |  |  | 10, 35, 50, 90 | 50, 90, 50, 10 |  |  | 50, 90, 50, 10 |  |

**Supplementary Table 3.** EWAS atlas enrichment analysis results for all CpGs selected into the DNA methylation sparse Partial Least Squares Discriminant Analysis (sPLS-DA) model. Enriched traits based on enrichment analysis with 486 CpGs selected by the 5-component DNA methylation sPLS-DA model. The fourth column (DMC) shows how many of the 486 CpGs have been previously associated with the trait in the first column. The fifth column (background) shows how many CpGs have previously been associated with the trait in column 1. The last column (%) shows the percentage of CpGs previously associated with the trait in column 1 that were also selected by the DNA methylation sPLS-DA model.

| **Trait** | **OR** | ***p*** | **DMC** | **Background** | **%** |
| --- | --- | --- | --- | --- | --- |
| glucocorticoid exposure | 7.66 | 1.12E-14 | 13 | 3468 | 0.37 |
| ancestry | 2.61 | 5.73E-06 | 15 | 10618 | 0.14 |
| Trihalomethanes (THM) exposure | 29.04 | 1.47E-05 | 2 | 140 | 1.43 |
| childhood stress | 11.00 | 2.46E-05 | 3 | 550 | 0.55 |
| household socioeconomic status in childhood | 9.75 | 4.75E-05 | 3 | 620 | 0.48 |
| ankylosing spondylitis | 18.21 | 8.64E-05 | 2 | 222 | 0.90 |
| osteonecrosis of the femoral head (ONFH) | 66.63 | 4.88E-04 | 1 | 31 | 3.23 |
| recurrent stroke | 24.09 | 3.39E-03 | 1 | 84 | 1.19 |
| primary Sjögren's Syndrome (pSS) | 2.84 | 3.64E-03 | 5 | 3526 | 0.14 |
| infertility | 2.27 | 8.89E-03 | 6 | 5281 | 0.11 |
| thyroid lesion | 3.43 | 9.40E-03 | 3 | 1753 | 0.17 |
| aging | 1.41 | 9.86E-03 | 23 | 31184 | 0.07 |

**Supplementary Table 4.** Prediction parameters of the sparse Partial Least Squares Discriminant Analyses (sPLS-DA) models in the test and clinical follow-up data. Balanced error rates (BER), prediction sensitivity, specificity, and accuracy, and model Area Under the Curve (AUC) of the prediction of the sPLS-DA models in the test and follow-up data per component. The AUC *p*-values have been adjusted separately for the test and follow-up data for multiple testing using the FDR of 5% for 5 tests (*q*).

| **Trait** | **Component** | **BER** | **Sensitivity** | **Specificity** | **Accuracy** | **AUC** | **AUC *p*** | **AUC *q*** |
| --- | --- | --- | --- | --- | --- | --- | --- | --- |
| **Test data** | | | | | | | | |
| Polygenic scores | 1 | 0.40 | 64.9 | 55.7 | 57.0 | 0.60 | 0.06 | 0.27 |
| Polygenic scores | 2 | 0.43 | 54.1 | 60.6 | 59.7 | 0.59 | 0.07 | 0.27 |
| DNA methylation | 1 | 0.46 | 37.8 | 69.2 | 64.7 | 0.51 | 0.80 | 0.81 |
| DNA methylation | 2 | 0.48 | 32.4 | 72.4 | 66.7 | 0.53 | 0.52 | 0.77 |
| DNA methylation | 3 | 0.45 | 29.7 | 80.5 | 73.3 | 0.52 | 0.64 | 0.77 |
| DNA methylation | 4 | 0.46 | 24.3 | 83.7 | 75.2 | 0.56 | 0.28 | 0.77 |
| DNA methylation | 5 | 0.46 | 21.6 | 87.3 | 77.9 | 0.60 | 0.05 | 0.27 |
| Metabolomics | 1 | 0.51 | 43.2 | 55.2 | 53.5 | 0.48 | 0.64 | 0.77 |
| Metabolomics | 2 | 0.55 | 29.7 | 59.7 | 55.4 | 0.47 | 0.52 | 0.77 |
| Metabolomics | 3 | 0.52 | 29.7 | 66.1 | 60.9 | 0.45 | 0.37 | 0.77 |
| Metabolomics | 4 | 0.50 | 32.4 | 67.4 | 62.4 | 0.46 | 0.41 | 0.77 |
| Metabolomics | 5 | 0.51 | 32.4 | 66.5 | 61.6 | 0.49 | 0.81 | 0.81 |
| **Clinical data** | | | | | | | | |
| Polygenic scores | 1 | 0.50 | 44.6 | 54.9 | 49.7 | 0.51 | 0.87 | 0.98 |
| Polygenic scores | 2 | 0.50 | 31.1 | 69.0 | 49.7 | 0.51 | 0.76 | 0.98 |
| DNA methylation | 1 | 0.57 | 23.0 | 63.4 | 42.8 | 0.46 | 0.44 | 0.98 |
| DNA methylation | 2 | 0.51 | 28.4 | 70.4 | 49.0 | 0.51 | 0.82 | 0.98 |
| DNA methylation | 3 | 0.48 | 32.4 | 71.8 | 51.7 | 0.51 | 0.76 | 0.98 |
| DNA methylation | 4 | 0.48 | 27.0 | 77.5 | 51.7 | 0.52 | 0.67 | 0.98 |
| DNA methylation | 5 | 0.46 | 20.3 | 87.3 | 53.1 | 0.54 | 0.47 | 0.98 |
| Metabolomics | 1 | 0.55 | 35.1 | 54.9 | 44.8 | 0.48 | 0.71 | 0.98 |
| Metabolomics | 2 | 0.50 | 35.1 | 54.9 | 44.8 | 0.48 | 0.71 | 0.98 |
| Metabolomics | 3 | 0.51 | 39.2 | 59.2 | 49.0 | 0.47 | 0.52 | 0.98 |
| Metabolomics | 4 | 0.47 | 37.8 | 67.6 | 52.4 | 0.50 | 0.98 | 0.98 |
| Metabolomics | 5 | 0.45 | 43.2 | 66.2 | 54.5 | 0.50 | 0.92 | 0.98 |

**Supplementary Table 5.** EWAS atlas enrichment analysis results for the CpGs included in cluster 1 and 2 of the polygenic scores (PGSs)-DNA methylation and the DNA methylation-metabolomics Partial Least Squares (PLS) models. Enriched traits based on enrichment analysis with 218 CpGs that were included in cluster 1 and 251 CpGs that were included in cluster 2 of the 2-component PGSs-DNA methylation PLS model, and the 241 CpGs that were included in cluster 1 and 245 CpGs that were included in cluster 2 of the 5-component DNA methylation-metabolomics PLS model. Cluster assignment is provided in **Supplementary Data 2**. The fourth column (DMC) shows how many of the CpGs have been previously associated with the trait in the first column. The fifth column (background) shows how many CpGs have previously been associated with the trait in column 1. The last column (%) shows the percentage of CpGs previously associated with the trait in column 1 that were also included in cluster 1 or cluster 2 of the PGSs-DNA methylation or DNA methylation-metabolomics PLS models.

| **Trait** | **OR** | ***p*** | **DMC** | **Background** | **%** |
| --- | --- | --- | --- | --- | --- |
| **PGS-DNA methylation cluster 1** | | | | | |
| glucocorticoid exposure | 12.94 | 6.64E-31 | 13 | 3468 | 0.37 |
| Trihalomethanes (THM) exposure | 33.89 | 1.85E-11 | 2 | 140 | 1.43 |
| recurrent stroke | 27.37 | 1.61E-06 | 1 | 84 | 1.19 |
| vitamin B12 supplement | 8.49 | 1.03E-04 | 1 | 589 | 0.17 |
| infertility | 2.57 | 3.68E-03 | 5 | 5281 | 0.09 |
| **PGS-DNA methylation cluster 2** | | | | | |
| ancestry | 4.32 | 3.30E-09 | 13 | 10618 | 0.12 |
| childhood stress | 20.69 | 7.20E-07 | 3 | 550 | 0.55 |
| household socioeconomic status in childhood | 18.34 | 1.43E-06 | 3 | 620 | 0.48 |
| ankylosing spondylitis | 34.18 | 7.71E-06 | 2 | 222 | 0.90 |
| aging | 2.10 | 1.24E-05 | 17 | 31184 | 0.05 |
| osteonecrosis of the femoral head (ONFH) | 125.17 | 1.41E-04 | 1 | 31 | 3.23 |
| type 2 diabetes (T2D) | 3.34 | 1.22E-03 | 5 | 5633 | 0.09 |
| neurodevelopmental presentations and congenital anomalies (ND/CAs) | 8.79 | 1.30E-03 | 2 | 856 | 0.23 |
| cystic fibrosis | 21.65 | 4.12E-03 | 1 | 174 | 0.57 |
| Klinefelter syndrome | 21.05 | 4.35E-03 | 1 | 179 | 0.56 |
| Coffin–Siris syndrome (CSS) | 14.03 | 9.44E-03 | 1 | 268 | 0.37 |
| **DNA methylation-metabolomics cluster 1** | | | | | |
| glucocorticoid exposure | 12.08 | 7.63E-30 | 13 | 3468 | 0.37 |
| Trihalomethanes (THM) exposure | 31.79 | 3.22E-11 | 2 | 140 | 1.43 |
| recurrent stroke | 25.67 | 2.19E-06 | 1 | 84 | 1.19 |
| vitamin B12 supplement | 7.96 | 1.45E-04 | 1 | 589 | 0.17 |
| infertility | 2.41 | 6.00E-03 | 5 | 5281 | 0.09 |
| **DNA methylation-metabolomics cluster 2** | | | | | |
| ancestry | 4.60 | 9.96E-10 | 13 | 10618 | 0.12 |
| childhood stress | 21.97 | 5.12E-07 | 3 | 550 | 0.55 |
| household socioeconomic status in childhood | 19.47 | 1.02E-06 | 3 | 620 | 0.48 |
| ankylosing spondylitis | 36.30 | 6.11E-06 | 2 | 222 | 0.90 |
| aging | 2.14 | 1.33E-05 | 16 | 31184 | 0.05 |
| osteonecrosis of the femoral head (ONFH) | 132.46 | 1.26E-04 | 1 | 31 | 3.23 |
| type 2 diabetes (T2D) | 3.55 | 7.85E-04 | 5 | 5633 | 0.09 |
| neurodevelopmental presentations and congenital anomalies (ND/CAs) | 9.34 | 1.04E-03 | 2 | 856 | 0.23 |
| cystic fibrosis | 22.98 | 3.67E-03 | 1 | 174 | 0.57 |
| Klinefelter syndrome | 22.34 | 3.88E-03 | 1 | 179 | 0.56 |
| Coffin–Siris syndrome (CSS) | 14.89 | 8.43E-03 | 1 | 268 | 0.37 |

**Supplementary Table 6.** EWAS atlas enrichment analysis results for all CpGs selected into the multi-block sparse Partial Least Squares Discriminant Analysis (MB-sPLS-DA) multi-omics model. Enriched traits based on enrichment analysis with 143 CpGs selected by the 4-component MB-sPLS-DA model. The fourth column (DMC) shows how many of the 143 CpGs have been previously associated with the trait in the first column. The fifth column (background) shows how many CpGs have previously been associated with the trait in column 1. The last column (%) shows the percentage of CpGs previously associated with the trait in column 1 that were also selected by the MB- sPLS-DA model.

| **Trait** | **OR** | ***p*** | **DMC** | **Background** | **%** |
| --- | --- | --- | --- | --- | --- |
| childhood stress | 38.00 | 2.25E-08 | 3 | 550 | 0.55 |
| respiratory allergies (RA) | 28.48 | 1.58E-05 | 2 | 485 | 0.41 |
| household socioeconomic status in childhood | 22.25 | 4.08E-05 | 2 | 620 | 0.32 |
| ancestry | 3.75 | 5.67E-05 | 7 | 10618 | 0.07 |
| colorectal laterally spreading tumor | 4.20 | 2.43E-04 | 5 | 8245 | 0.06 |
| follicular thyroid carcinoma | 4.96 | 3.29E-04 | 4 | 5575 | 0.07 |
| type 2 diabetes (T2D) | 4.91 | 3.52E-04 | 4 | 5633 | 0.07 |
| aging | 2.13 | 8.01E-04 | 11 | 31184 | 0.04 |
| response to antidepressants | 49.56 | 8.27E-04 | 1 | 139 | 0.72 |
| short-term diesel exhaust inhalation | 47.51 | 8.99E-04 | 1 | 145 | 0.69 |
| multiple system atrophy | 43.81 | 1.05E-03 | 1 | 157 | 0.64 |
| Parkinson's disease (PD) | 24.65 | 3.21E-03 | 1 | 278 | 0.36 |
| obesity | 3.38 | 3.55E-03 | 4 | 8134 | 0.05 |

**Supplementary Table 7.** Prediction parameters of the multi-block sparse Partial Least Squares Discriminant Analyses (MB-sPLS-DA) model in the test and clinical follow-up data. Balanced error rates (BER), prediction sensitivity, specificity, and accuracy, and model Area Under the Curve (AUC) of the prediction of the MB-sPLS-DA model in the test and follow-up data per component. The AUCs are reported for the polygenic score, DNA methylation, and metabolomics levels included in the MB-sPLS-DA model, separately. The AUC *p*-values have been adjusted separately for the test and follow-up data for multiple testing using the FDR of 5% for 48 tests (*q*).

|  |  |  |  |  | **polygenic scores** | | | **DNA methylation** | | | **Metabolomics** | | |
| --- | --- | --- | --- | --- | --- | --- | --- | --- | --- | --- | --- | --- | --- |
| **Component** | **BER** | **Sensitivity** | **Specificity** | **Accuracy** | **AUC** | ***p*** | ***q*** | **AUC** | ***p*** | ***q*** | **AUC** | ***p*** | ***q*** |
| **Test data** | | | | | | | | | | | | | |
| 1 | 0.42 | 54.1 | 61.5 | 60.5 | 0.62 | 0.02 | 0.08 | 0.5 | 0.94 | 0.94 | 0.46 | 0.41 | 0.68 |
| 2 | 0.51 | 35.1 | 62.9 | 58.9 | 0.54 | 0.48 | 0.68 | 0.48 | 0.73 | 0.84 | 0.47 | 0.56 | 0.68 |
| 3 | 0.49 | 29.7 | 71.5 | 65.5 | 0.53 | 0.5 | 0.68 | 0.51 | 0.85 | 0.91 | 0.44 | 0.23 | 0.52 |
| 4 | 0.5 | 27 | 72.4 | 65.9 | 0.53 | 0.51 | 0.68 | 0.54 | 0.48 | 0.68 | 0.43 | 0.16 | 0.43 |
| **Clinical Data** | | | | | | | | | | | | | |
| 1 | 0.51 | 36.5 | 62 | 49 | 0.52 | 0.69 | 0.95 | 0.44 | 0.22 | 0.59 | 0.52 | 0.72 | 0.95 |
| 2 | 0.49 | 29.7 | 71.8 | 50.3 | 0.51 | 0.88 | 0.97 | 0.48 | 0.68 | 0.95 | 0.53 | 0.5 | 0.95 |
| 3 | 0.55 | 25.7 | 64.8 | 44.8 | 0.51 | 0.91 | 0.97 | 0.44 | 0.22 | 0.59 | 0.51 | 0.78 | 0.96 |
| 4 | 0.54 | 27 | 64.8 | 45.5 | 0.5 | 0.97 | 0.97 | 0.46 | 0.36 | 0.82 | 0.52 | 0.7 | 0.95 |
